# Violence against women and children in Yemen: A mixed-methods systematic review

**DOI:** 10.1101/2024.08.01.24310001

**Authors:** Samana Shreedhar, Sayali Arvind Chavan, Marwah Al-Zumair, Mirna Naccache, Priya Shreedhar, Lauren Maxwell

## Abstract

Violence against women and children (VAWC) is a significant health and human rights issue closely tied to multiple Sustainable Development Goals. While VAWC is prevalent in all countries, the severity and incidence of VAWC increase during wars, natural disasters, economic crises, and pandemics, all of which have affected Yemen in recent years. This systematic review synthesizes evidence from qualitative and quantitative studies on the types, prevalence, perpetrators of, and risk factors for VAWC in Yemen. Before initiating the search, the protocol and search strategy were registered to PROSPERO (CRD42021237855). We systematically searched four biomedical databases and grey literature sources and used reverse snowball sampling to identify eligible studies. The 31 studies included in the analysis depicted a range of forms of VAWC, encompassing honor violence, female genital mutilation and cutting, early and very early marriage, tourist marriage, family and intimate partner violence, and gender inequities in access to food, education, and medical care. Included studies reported a high prevalence of many forms of violence, including corporal punishment in schools and intimate partner violence. We reviewed study quality and how studies addressed ethical concerns in VAWC-related research. We found that several studies did not report ethics review or interviewer training and no studies discussed safety planning or addressing the mental health needs of participants in VAWC research. This systematic review provides a much-needed synthesis of existing research on VAWC in Yemen. Since the start of the 2014 war, Yemen has become the world’s largest humanitarian crisis, with the highest rate of maternal mortality and gender inequality in the world. We only identified one study initiated after the recent war in Yemen. This deficiency represents a missed opportunity to understand how the ongoing war has reversed prior gains in reducing the prevalence of child and very early child marriage and introduced new forms of gender-based violence, including tourist marriage.

## Introduction

Violence against women and children (VAWC) is a global human rights issue that closely relates to the achievement of the Sustainable Development Goals (SDGs), particularly Goal 5, which aims to achieve gender equality and empower all women and girls and Goal 16, which seeks to promote peaceful and inclusive societies for sustainable development, provide access to justice for all, and build effective, accountable institutions. In this systematic review (SR), we summarize primary and secondary research related to the forms, perpetrators, consequences, and reporting of VAWC in Yemen to inform future research and the development of interventions that prevent VAWC in Yemen or address the consequences of VAWC.

### Humanitarian crisis

In 2023, eighty percent of Yemen’s population of 29.8 million people required humanitarian assistance [1], and 4.5 million people were internally displaced [2], making Yemen the world’s largest humanitarian crisis. The recent war has decimated the country’s economy, healthcare, education, and infrastructure [3]. In addition to the ongoing humanitarian crisis, Yemen has deeply entrenched gender inequities where limited resources mean that women and girls do not have the same access to limited educational, health care, or food resources as their male counterparts [4,5]. As in any humanitarian crisis [6], the prolonged war disproportionately affected Yemeni women and children.

Non-governmental organizations (NGOs) in Yemen report increased levels of forced, early marriage and trafficking of girls and young children in response to the ongoing war [7]. Girls are sexually exploited in bars and restaurants in the larger towns of Yemen and may be forced to traffic illegal drugs [8]. Both Saudi and Houthi forces have used child soldiers [9]. The economic crisis and targeted attacks on infrastructure, including education, health care, and government services, mean that child labor has become commonplace [8], affecting mental and physical health (19). Where school bombings have not occurred, parents are reluctant to send their children to school for fear of bombing, trafficking, recruitment of child soldiers, and other forms of community or war-related violence [10].

### Violence against women

Intimate partner violence (IPV), defined as “behavior by an intimate partner or ex-partner that leads to physical, sexual or psychological harm, including aggression, sexual coercion, psychological abuse, and controlling behaviors” [11], is the most prevalent form of gender-based violence (GBV) globally. Longstanding gender inequities codified into law mean that Yemeni women, their families, and their community may perceive their experience of IPV, including marital rape, as their fault [12]. Societal stigma associated with physical and sexual violence means that women and children have limited options for support or for reporting violence. Where support is available, the stigmatization of women or children who experience violence, especially sexual violence, prevents reporting or accessing available services [13].

### Violence against children

The World Health Organization (WHO) defines violence against children (VAC) as ‘’violence in all forms experienced by children who are under 18 years old, perpetrated by parents or other caregivers, peers, romantic partners, or strangers” [14,15]. Globally, 75% of children experience physical or emotional abuse from their parents, peers, or caretakers [14,15]. Female genital mutilation or cutting (FGM/C), defined as “all procedures involving partial or total removal of the external female genitalia or other intentional injuries to the female genital organs for non-medical reasons,” [16] is a prevalent cultural practice in Yemen and elsewhere in the Middle East and North Africa (MENA) and sub-Saharan Africa (SSA) [17]. The practice of FGM/C leads to an increase in urinary, vaginal, and pregnancy-related morbidities and mortalities, repeat surgeries, and death [12].

#### Gender inequitable access to food, education, employment, health care, and legal protections

In 2021, Yemen ranked last out of 170 countries on the Gender Inequality Index (GII) [18]. Educational attainment and literacy in Yemen are low (8, 10), especially for women and only 6% of women work outside of their home [5]. Yemen has two legal systems, *Shari’a* and *Kabeelee,* or tribal law. Under *Shari’a* law, Yemeni women lack child custody protections and have limited to no right to divorce, property rights, or the right to manage their finances [19]. In contrast, men are legally entitled to control their spouse’s money and property, are the default recipients of child custody, and can divorce their wives by saying “talaq’’ [19]. Marital rape remains non-criminalized [20] even though Yemen acceded to the Convention on the Elimination of All Forms of Discrimination Against Women (CEDAW) in 1984, mandating states to ensure that marriage is founded on free and full consent [21]. Yemen does not have a minimum age of marriage, even though Yemen ratified the Convention on the Rights of the Child in 1989, setting 18 as the minimum age for marriage.

#### Intersectionality and violence against women and children

Minority ethnic groups in Yemen, including the indigenous *Akhdam* or *Muhamasheen* community and refugees from Somalia, Sudan, or Ethiopia, face high levels of discrimination and violence in Yemen, including sexual assault and rape [22]. Homosexuality in Yemen is a criminal offence under *Shari’a* law that is punishable by death, which means that violence against lesbian, gay, bisexual, transgender, queer, and intersex (LGBTQI) likely goes unreported [22].

## Materials and Methods

We used a mixed-methods SR design with a narrative synthesis approach to provide a broad overview of VAWC that could be used to inform policymakers and develop research priorities [23]. The SR protocol was developed using the 2015 Preferred Reporting Items for Systematic Reviews and Meta-Analyses Protocols (PRISMA-P) guidelines [24] and registered on the PROSPERO Register of Systematic Reviews before initiating the search (CRD42021237855). We followed the 2020 PRISMA guidelines for conducting and reporting systematic reviews [25] and reported the PRISMA elements in Supplementary Table S1.

### Search strategy

We conducted a systematic search of four electronic databases, Ovid (MEDLINE) and (Embase), Cumulated Index to Nursing and Allied Health Literature (CINAHL), and Web of Science (WoS), using a combination of MeSH and text terms (S2 Table). We ran the searches on 18 February 2021 and did not include language or date restrictions. We updated the search for Ovid (Medline), WoS, and CINAHL on 8 April 2024 but no longer had access to Ovid (Embase) and did not rerun that search. We also searched for additional citations using backward snowball sampling [26], reviewing the citations in included studies. Additional studies were identified through grey literature searches, using the Google search engine and reviewing NGO websites, including the United Nations High Commissioner for Refugees (UNHCR), Save the Children, United Nations Children’s Fund (UNICEF), and United Nations Family Planning Association (UNFPA) websites. We also reviewed Yemen country reports from the Demographic and Health Surveys (DHS) and the Multi-Cluster Indicator Surveys (MICS), population-representative survey research initiatives that focus on maternal and child health (MCH)-related outcomes.

#### Inclusion criteria

Included studies were primary research studies that used any design and that reported on any form of sexual, physical, emotional, or economic violence against Yemeni women and children, including IPV and family violence (early or very early marriage or forced marriage, femicide, prenatal sex selection, female infanticide, FGM/C, gendered access to food, education, vaccines, healthcare, employment; dowry-related violence; honor killings; child labor; child neglect and maltreatment) workplace violence; war-related violence (child soldiers, captivity, trafficking of women and children, war-related sexual violence, physical attacks on women and children (e.g., rape, bombing), indirect attacks (bombing of water treatment facilities, hospitals, schools); pandemic-related violence (child labor, increased family, or IPV), violence related to displacement, gendered access to masks or COVID-19 treatments or vaccines. Studies that did not report on VAWC in Yemen and publications that did not represent primary research or secondary analyses of participant-level data (e.g., commentaries, systematic reviews, editorials) were excluded.

### Data extraction and management

Citations were exported to EndNote (version 9.3.3) for deduplication using the Braner approach [27]. Citations were then exported to Covidence version 2.0 [28] for title-abstract and full-text screening and data extraction using a pre-piloted data extraction form (Supplementary Text 5). Three reviewers (SC, MAL, SS) independently conducted the screening and data extraction. A fourth reviewer (LM) resolved disagreements. A meta-analysis was not possible due to the heterogeneity of study designs, types of violence, and age groups. We used thematic synthesis [29] to summarize findings across studies. We treated different types of violence as themes and identified additional themes, including risk factors, perpetrators, resources, and barriers to accessing resources, from the included studies and our review of related literature.

### Quality assessment

We assessed the quality of included primary research studies using the Mixed Method Analysis Tool (MMAT), version 2018 [30], which reviews the study’s design, sampling strategy, generalizability, data collection methods, integrity of intervention (where applicable), and analytic approach. We did not review the quality of population-level surveys (e.g., MICS, DHS, Arab Family Survey), which are generally regarded as high quality. Quality assessment criteria were applied by two independent reviewers (SS, MN); disagreements were resolved through a third reviewer (LM).

## Results

The screening process is summarized in the PRISMA Flow diagram [25] shown in Figure 1. From the initial search of the database conducted in February 2021, we identified 459 abstracts, while in the updated search done in April 2024, 106 abstracts were identified. After removing duplicates, 356 titles and abstracts (291 from the initial search and 65 from the updated search) were reviewed in Covidence. Subsequently, 63 studies were assessed for full-text eligibility, with 32 studies excluded for being systematic reviews, commentaries, editorials, or other non-primary research (19 studies) or for not measuring violence or stigma against Yemeni women and children (13 studies). Data was extracted from the 31 studies (6 reports and 25 articles(2 of which are grey literature sources)) that met the inclusion criteria. For details on the reasons for exclusion at the full-text review stage, refer to Supplementary Table S3.

### Overview of included studies

Table 1 provides an overview of the types of violence, risk factors, and consequences examined in the included studies, excluding data from the Yemen DHS and MICS reports (n=4) as they were included in other studies in the table. We identified 17 primary research studies, six reports, and eight secondary analyses that met the study inclusion criteria. Studies and reports included a diversity of age groups and types of violence and covered all of Yemen’s 22 governorates. Primary research included four qualitative studies and two qualitative components of mixed-method studies [7,21,31–34]. Most qualitative studies conducted in-depth interviews (IDIs; N=3), one study used a focus group discussion (FGD) approach [21], and two studies analyzed answers to open-ended questions in a cross-sectional questionnaire [32,34]. Supplementary Table S4 provides a detailed overview of quantitative studies and the quantitative estimates in mixed studies, including types of violence, measurement approach, point estimates (e.g., prevalence), measures of uncertainty (if any), associated risk factors, and regional differences. All 20 quantitative studies employed a cross-sectional design. Four quantitative studies [16,35–37] used DHS data [12,38,39]. Two studies [36,40] used data from MICSs [41], and five studies used data from various national and sub-national household surveys conducted in Yemen [42–46].

**Table 1.**
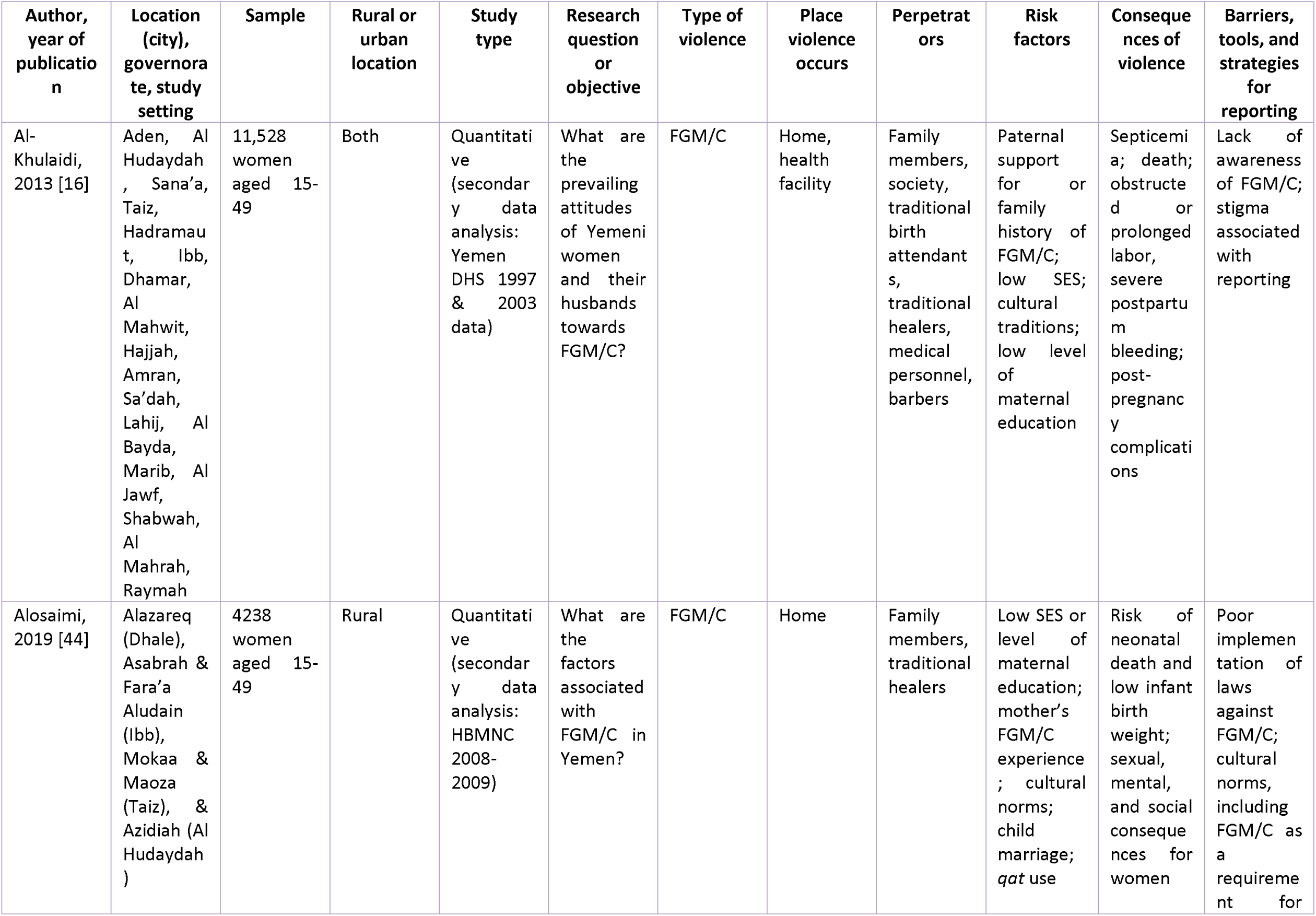
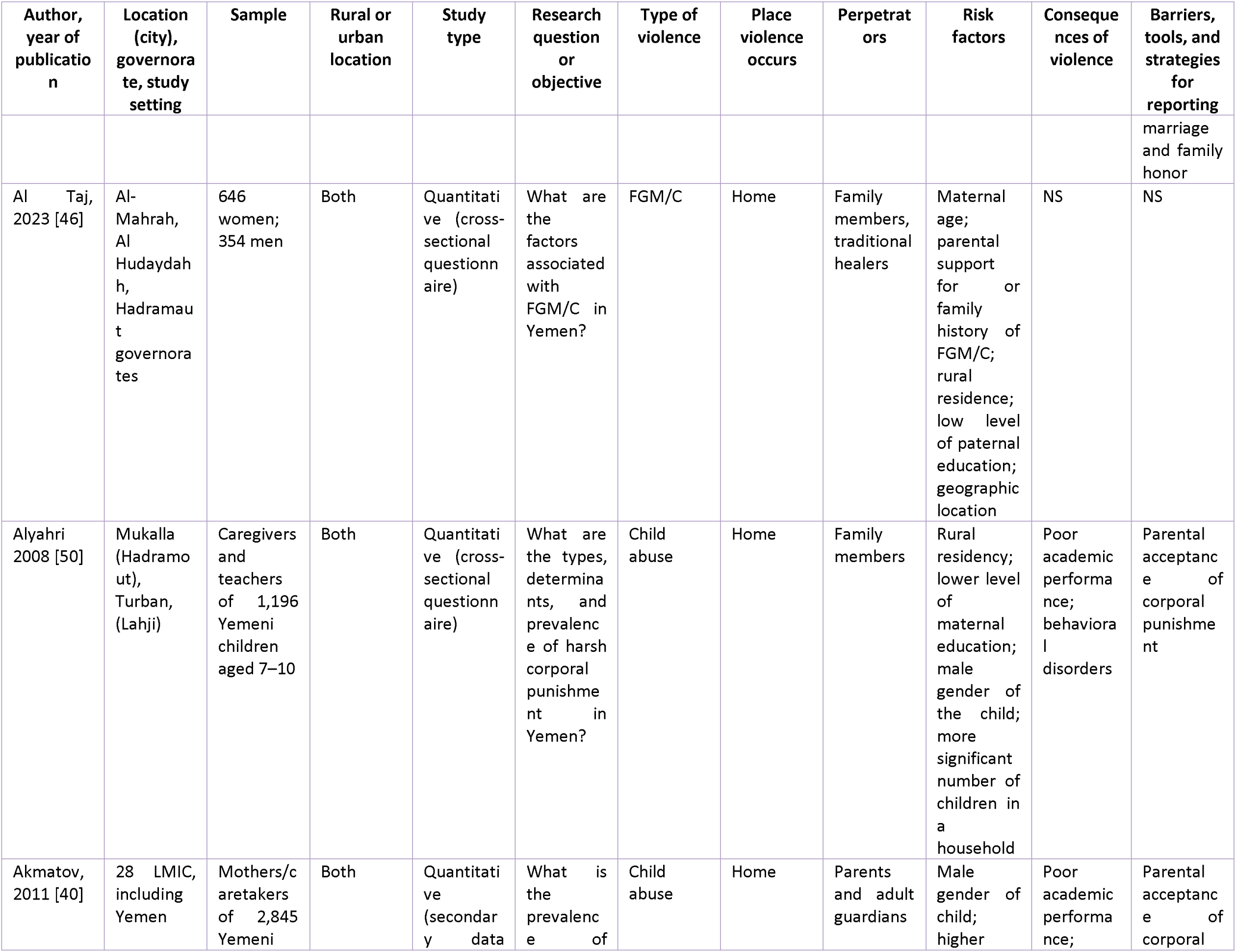
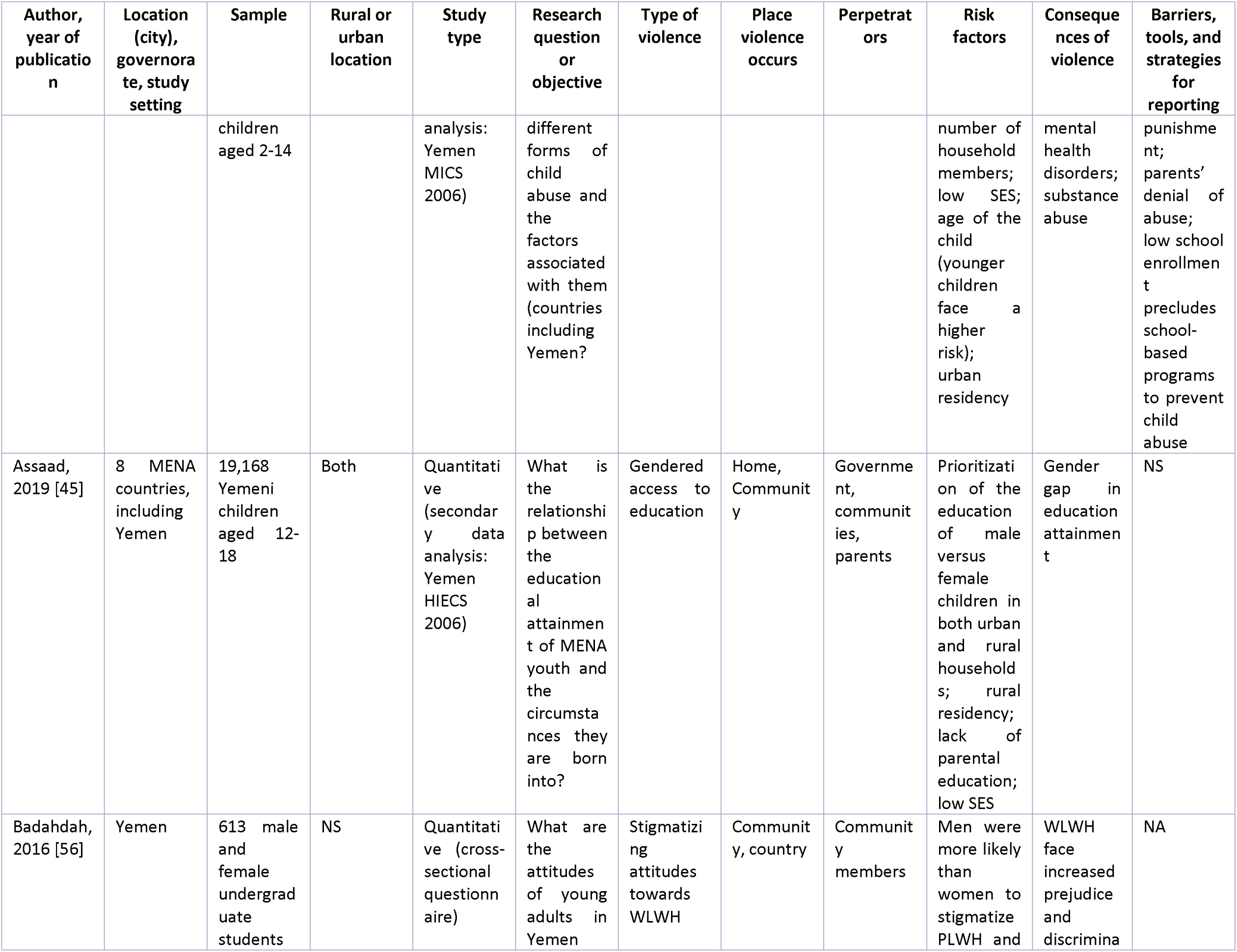
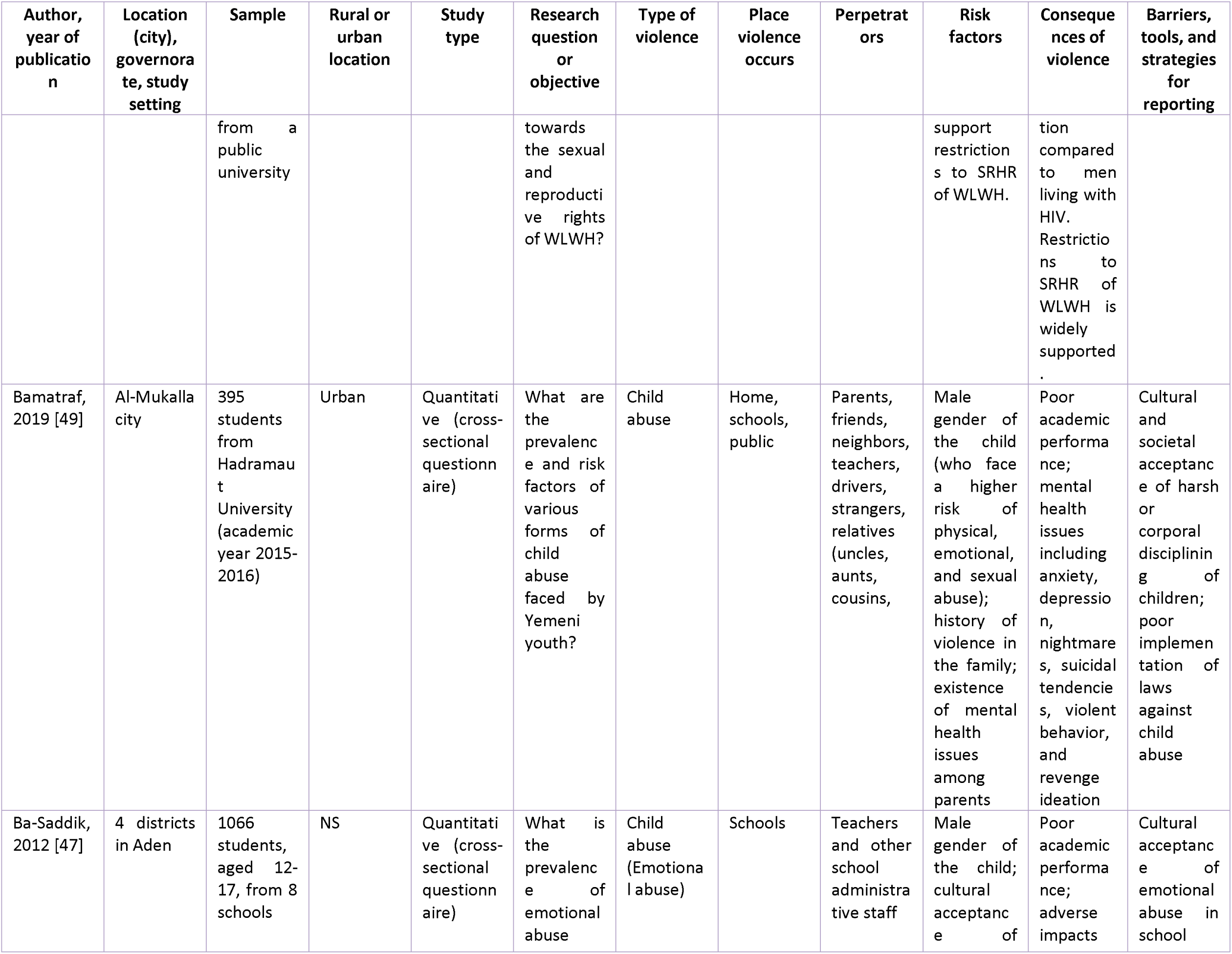
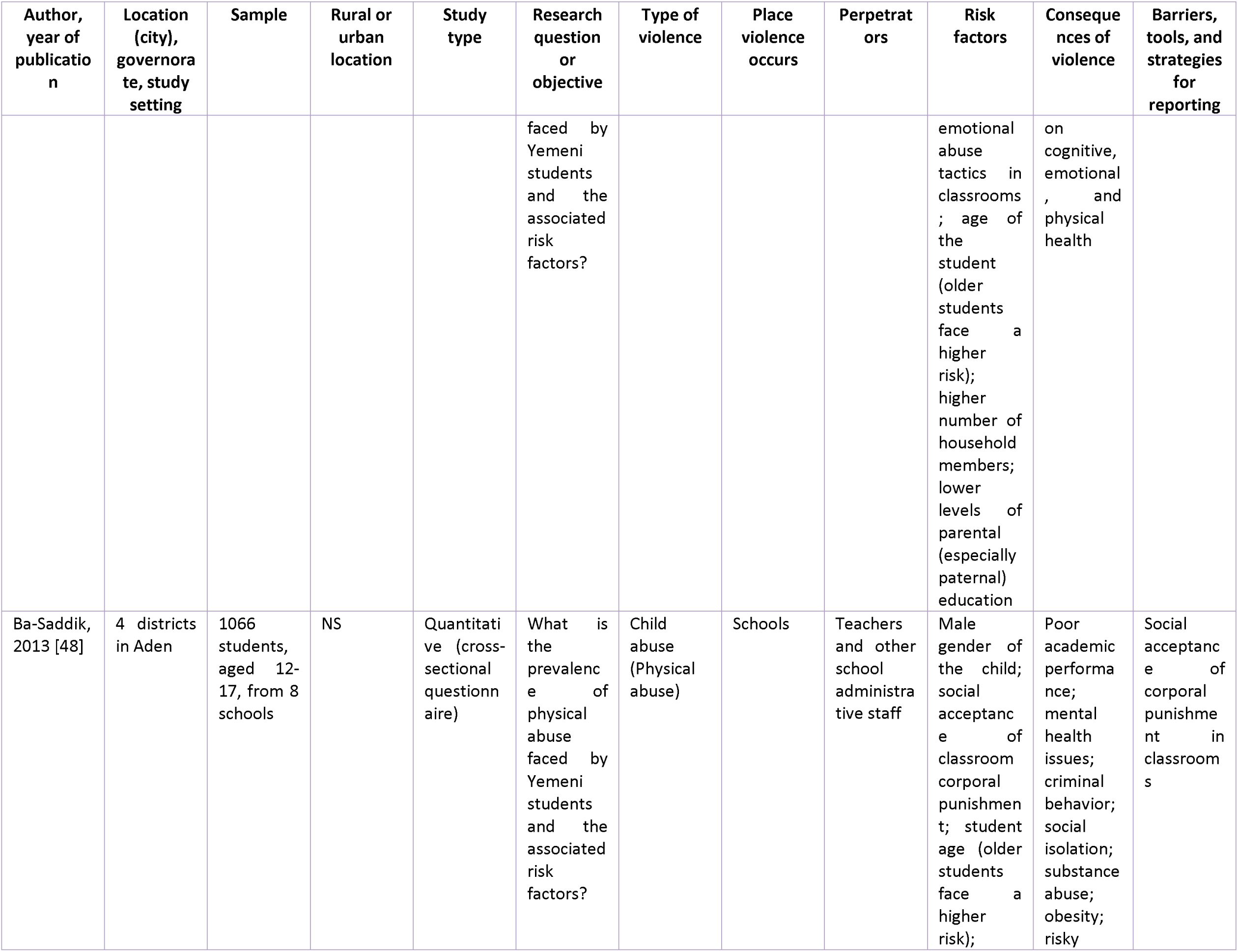
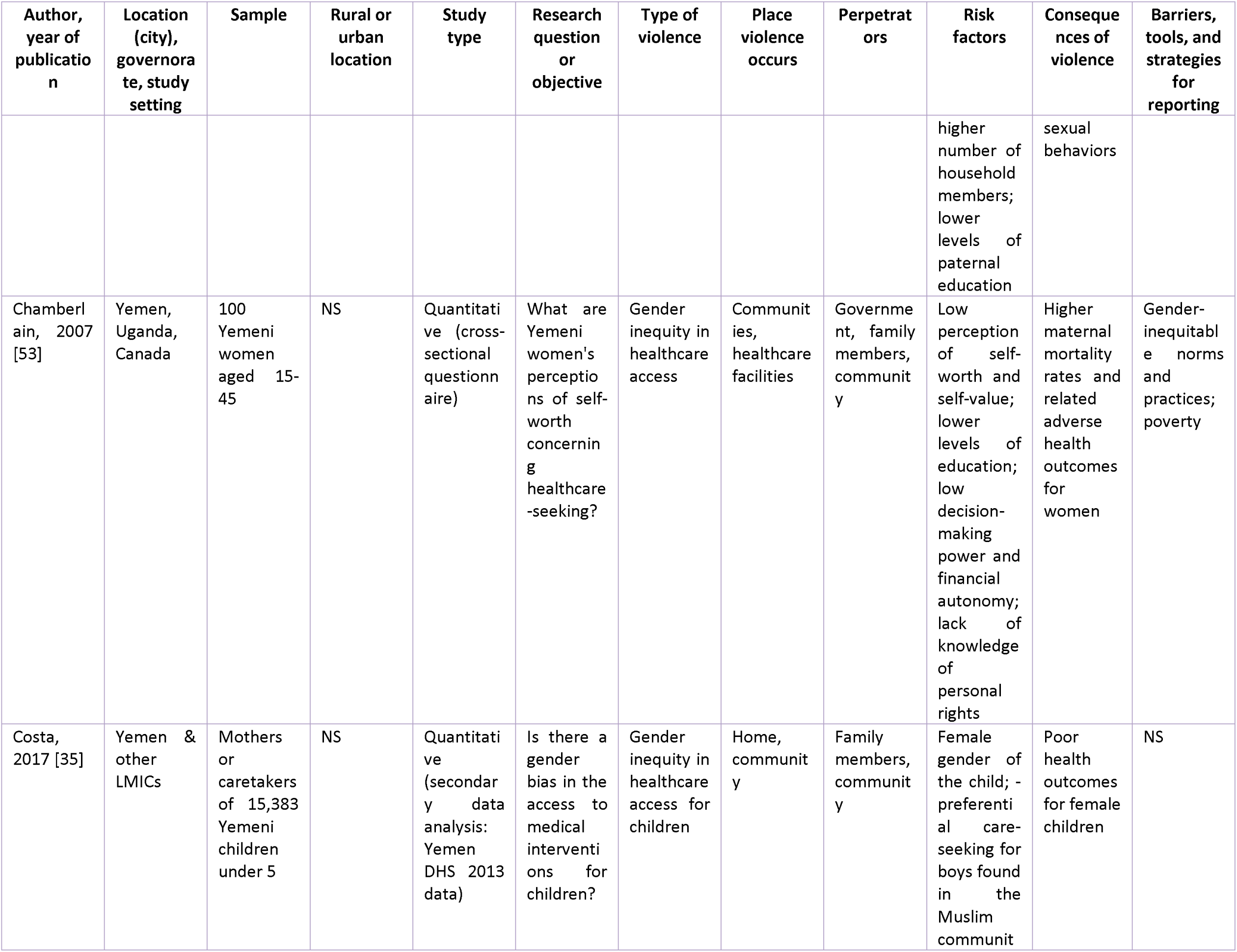
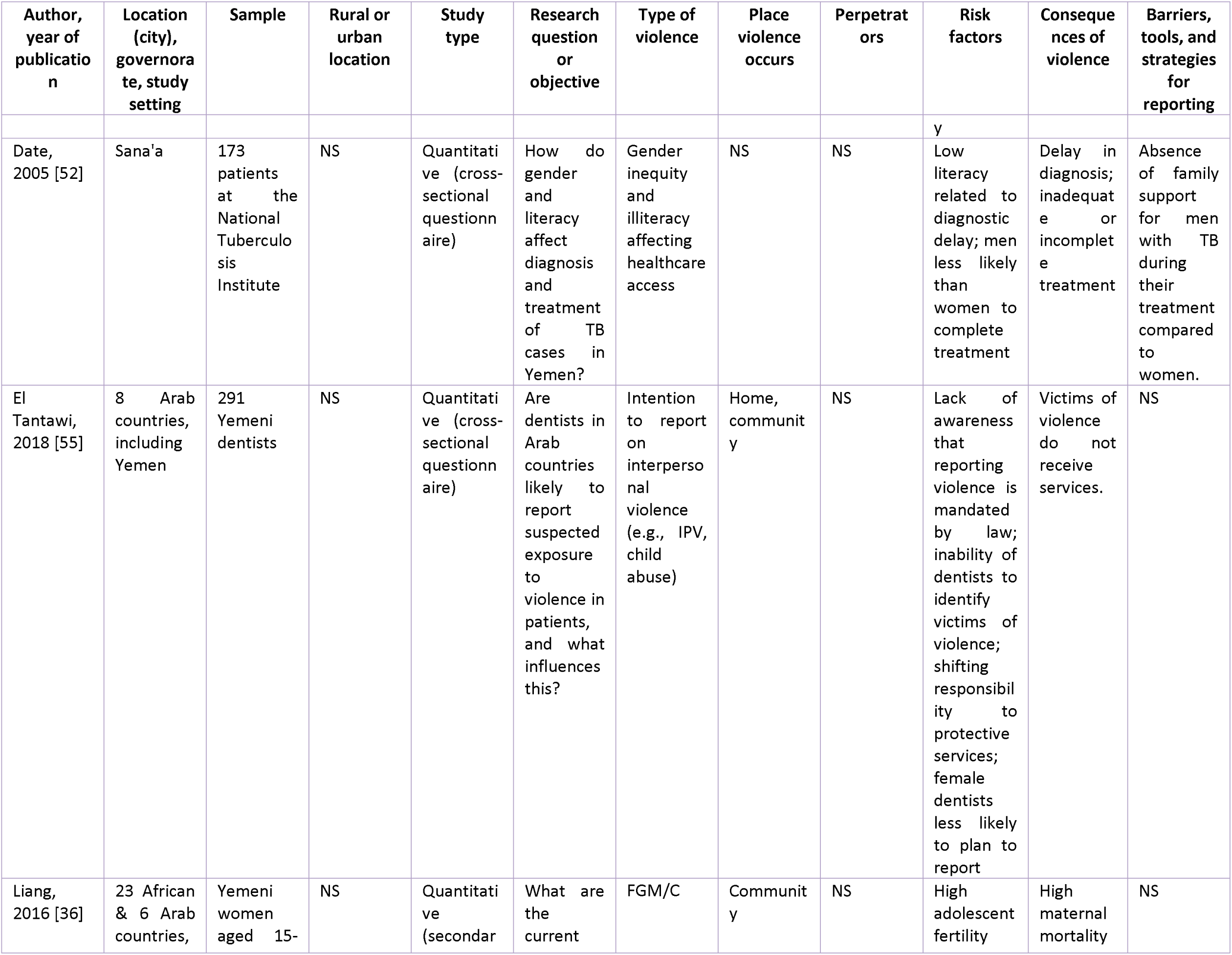
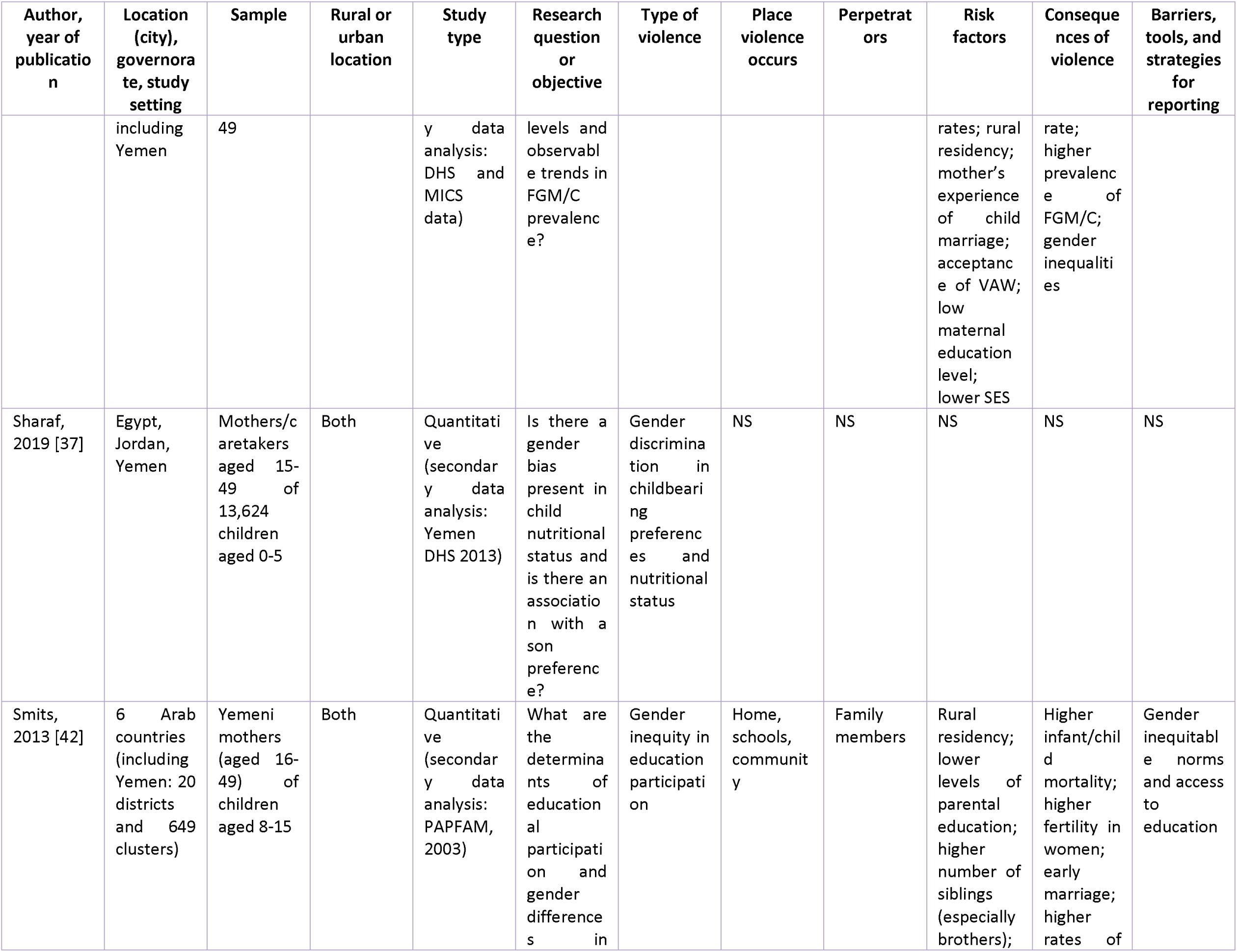
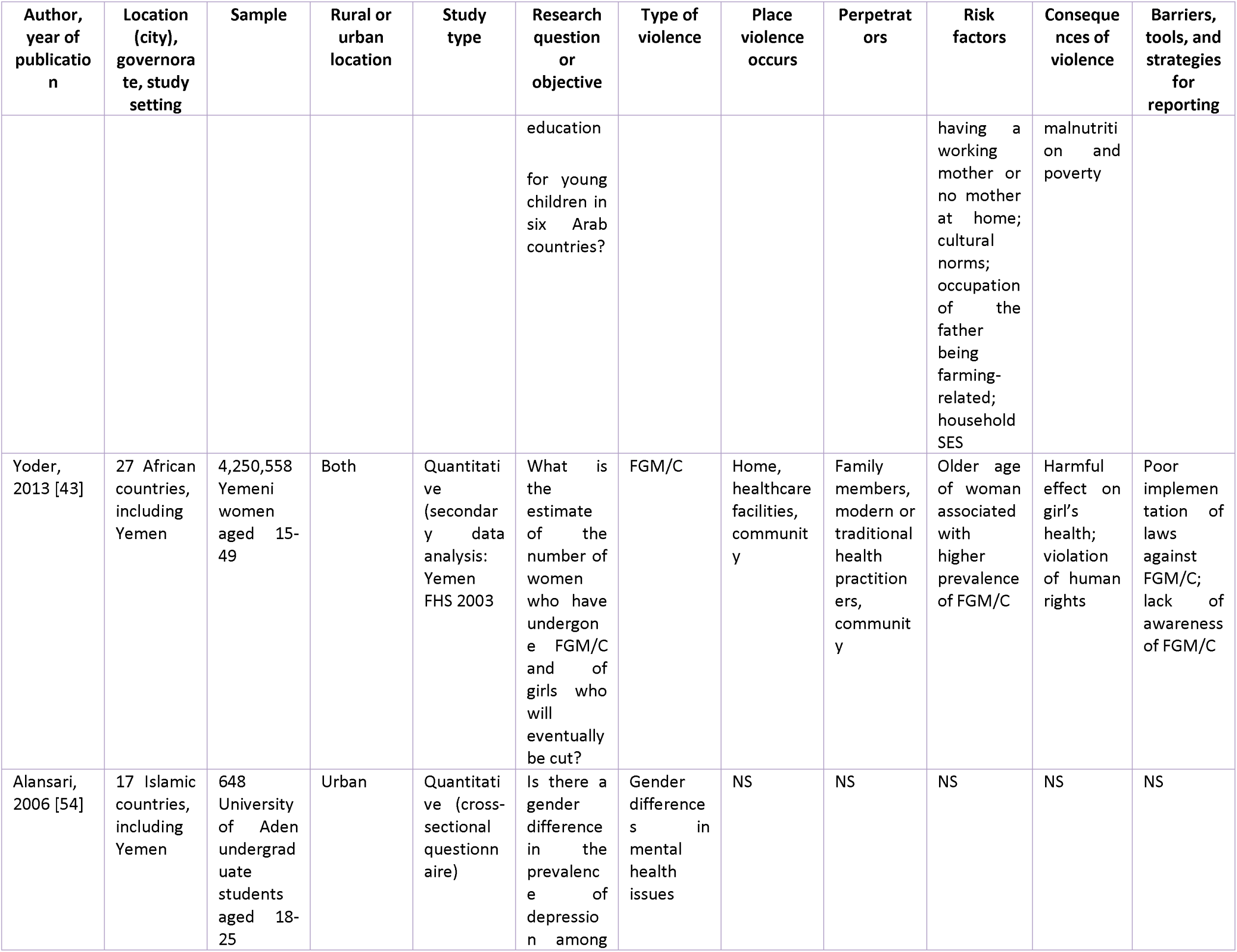
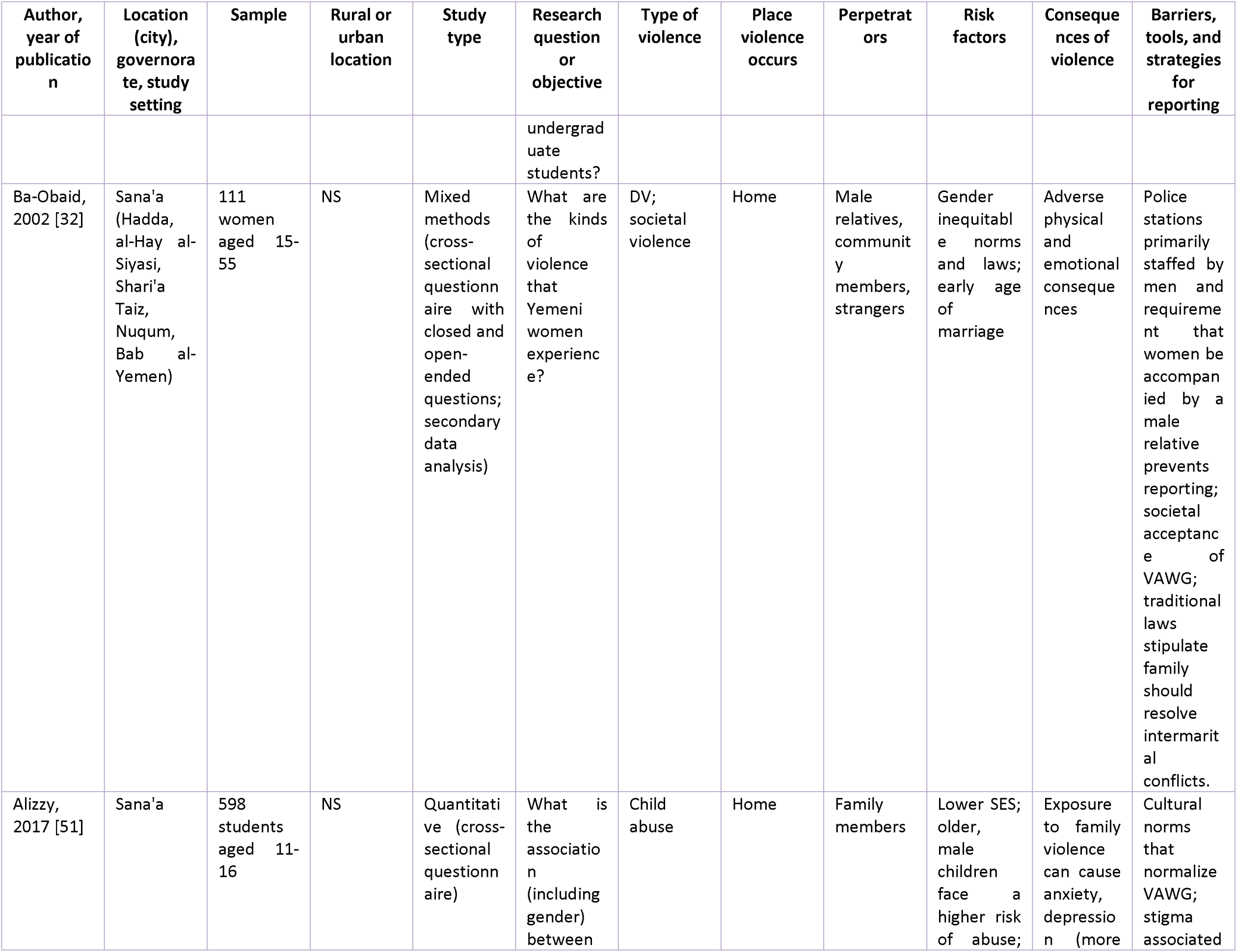
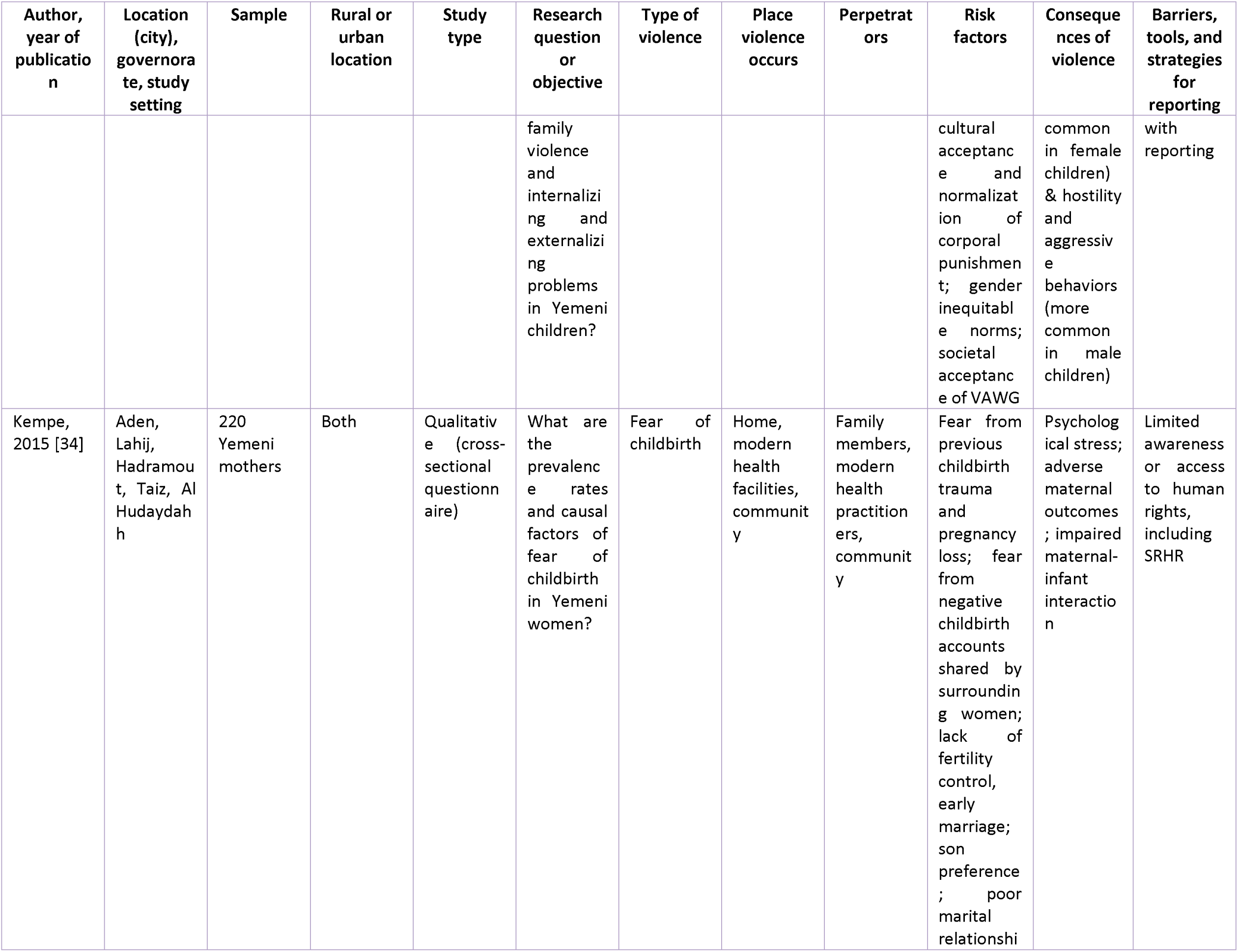
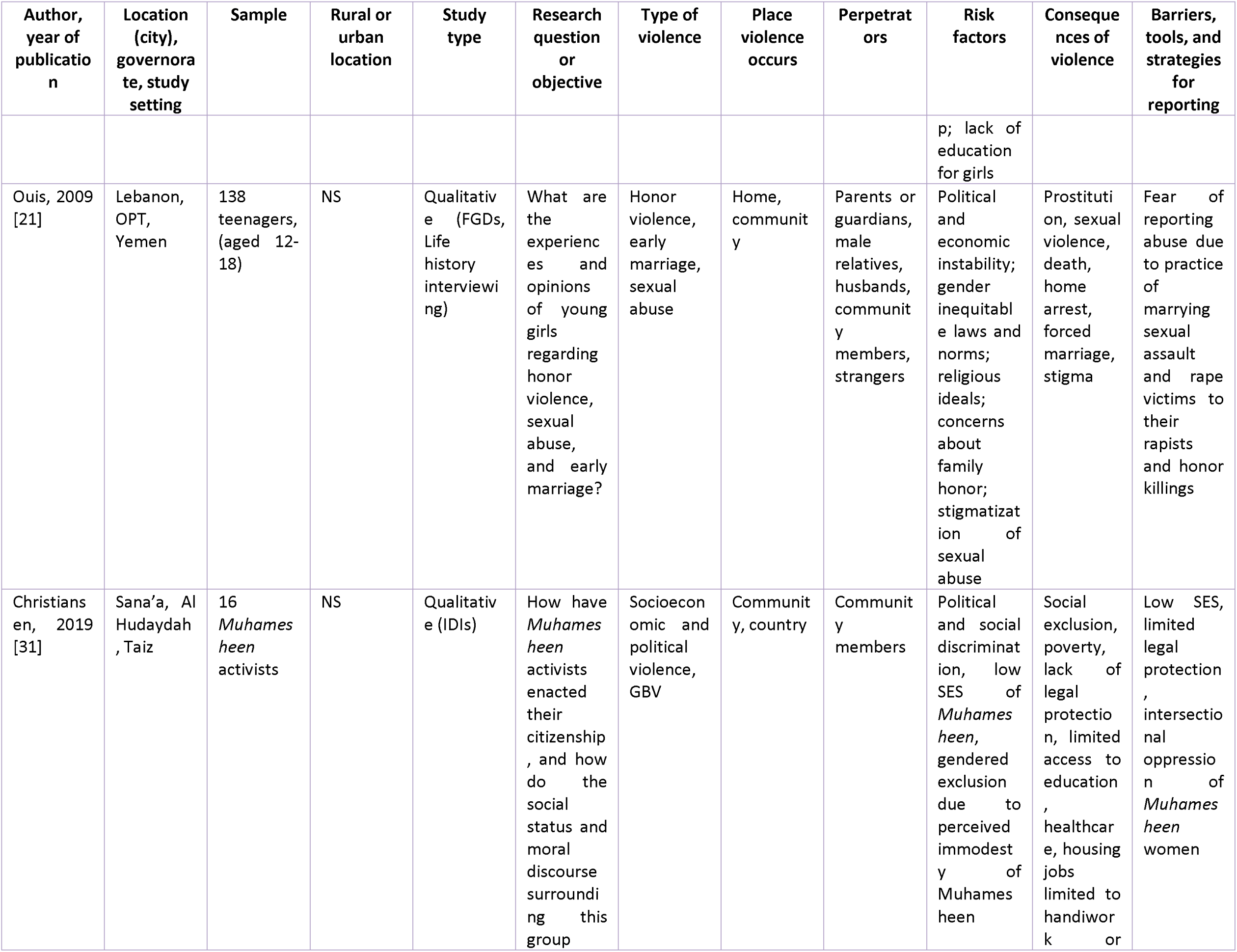
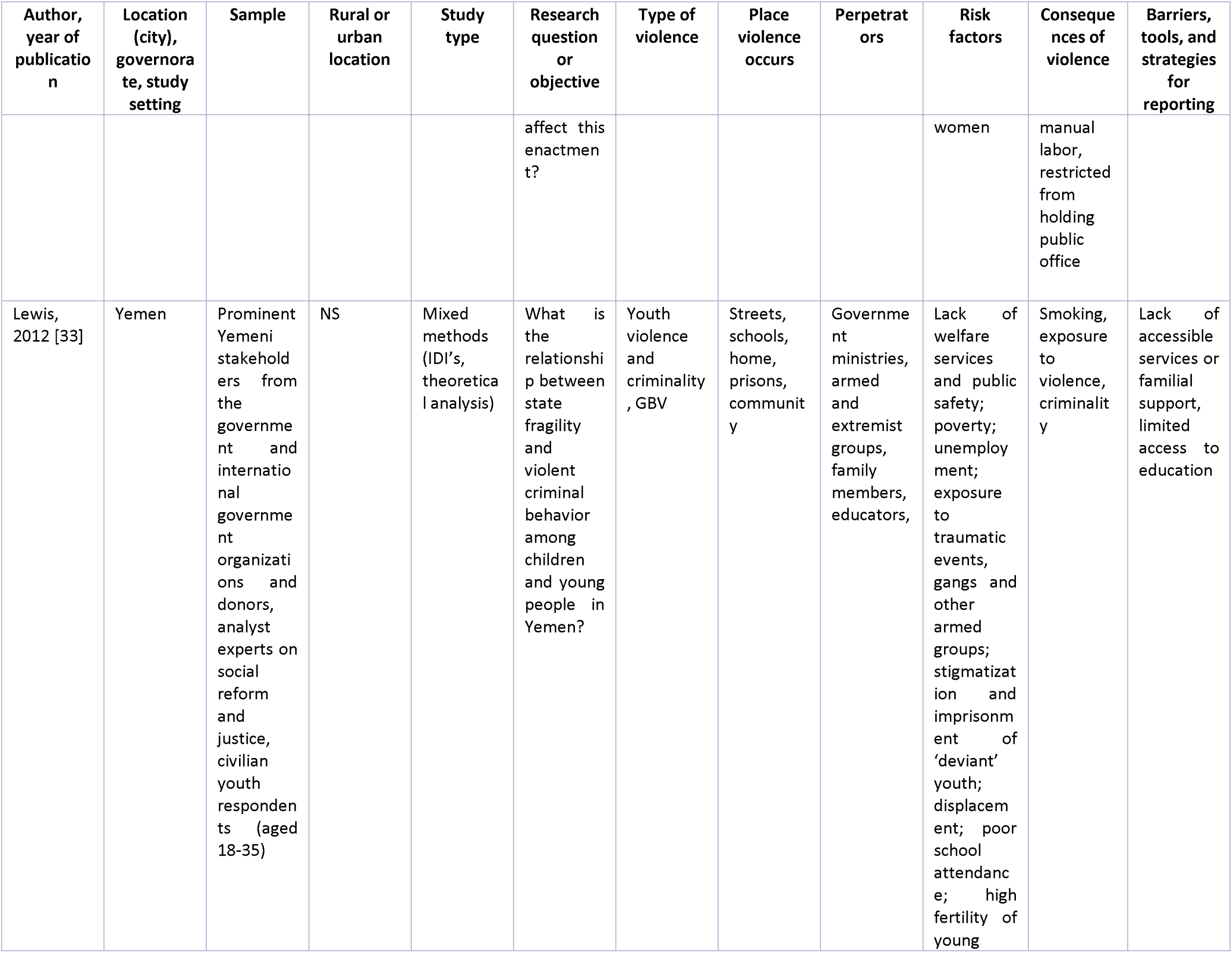
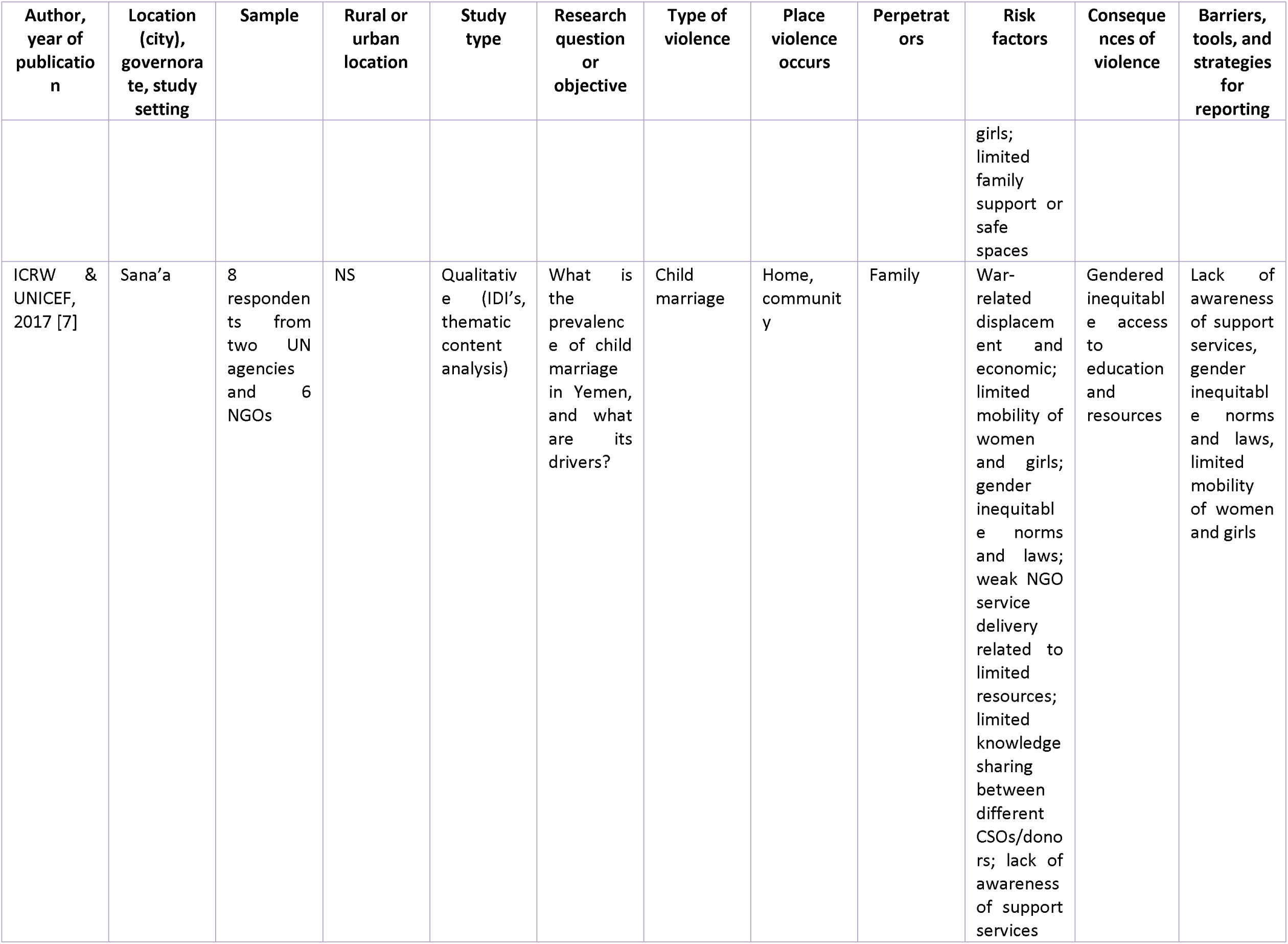
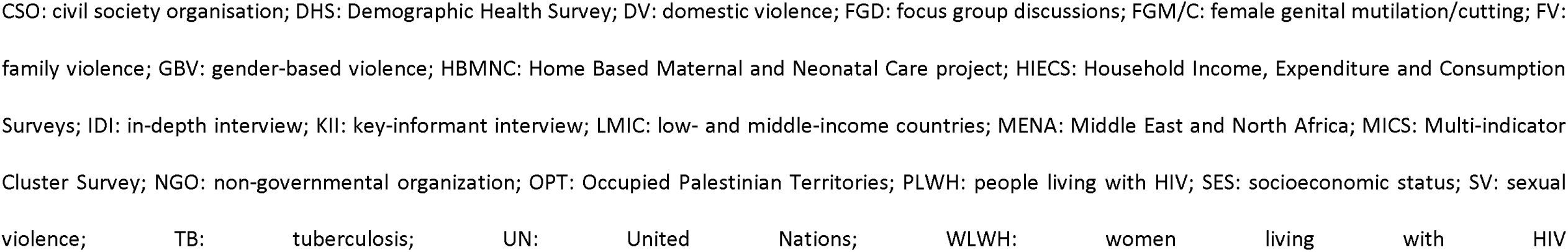
Overview of included studies.

Studies included different types of VAWC, offenders or perpetrators of violence, location of violence, risk factors, consequences of violence, and information on ethical concerns, access to services, or safety planning. We identified two studies related to early child marriage [7,21], seven studies on child abuse or maltreatment [21,40,47–51], five studies on FGM/C [16,36,43,44,46], seven studies on gender differences in education or health care services [35,37,42,45,52–54], and one study on community violence [31], IPV [32], attitudes towards honor violence [21], GBV [33], gender preference [37], dentist’s reports of suspected violence [55], and stigmatizing attitudes towards women with human immunodeficiency virus (HIV) [56]. No studies presented information on pandemic or war-related VAWC.

All studies reported on risk factors and perpetrators, and most studies described barriers to reporting violence [7,16,31–34,36,40,42–44,47–53,55,57]. Other than the DHS and MICS, which are country-wide, studies that collected primary data were conducted in Aden, Sana’a, Al Hudaydah, Taiz, Al-Mukalla, Al-Mahrah, Lahij, and Hadramout governorates. Most studies included both urban and rural areas, although a few studies were limited to urban (N=2) [49,54] or rural (N=1) [44] populations. In most studies, verbal consent, as opposed to written consent, was obtained. This practice could be attributed to concerns about participant safety in research related to VAWC or to Yemen’s generally low literacy level. Over half of the studies (25 of 31) were conducted before the 2014 war. While most VAWC occurs within the home, three articles found that women were also likely to experience GBV outside the home [32–34].

### Types of VAWC

#### Family and intimate partner violence against women

Neither the DHS from 1991/92, 1997, or 2013 nor the 2005/6 MICS included questions on women’s experience of partner or family violence. In addition to the DHS and MICS, two quantitative studies examined IPV [21,32]. One 2002 study of a stratified random sample of 111 ever-partnered women aged 15-55 in two areas of Sana’a found that 55% of women had experienced physical IPV (beatings, confinement to home, torture), 51% experienced threats, and 17% experienced sexual IPV [32]. Participants also reported experiencing violence perpetrated by their sons, brothers, fathers, mothers, and uncles [32]. Many participants reported that the violence they experienced was ‘’normal’’ and expected given societal norms [32]. Honor violence often occurs as a punitive response to perceived sexual immodesty in young girls, including after cases of sexual violence where the attribution of blame lies with the victim [21]. A 2009 qualitative study found that teenage girls in Yemen internalized honor violence. Participants reported acceptance of harsh punishments for girls who experienced sexual violence because they had ‘allowed’ or invited the violence [21].

#### Early and very early child marriage

The 1991/92 DHS revealed that 31% of girls were married by the age of 15 [38], a figure that decreased to 29% in the 1997 DHS [39] and further dropped to 14% in the 2006 MICS [41], and significantly decreased to 3% in the 2013 DHS [12]. A 2009 focus group study involving 138 Yemeni girls aged 12 to 18 discovered that financially insecure families felt compelled to marry off their young daughters [21].

> “First, I got married at 11, and second, my maternal uncle married me off. He found the groom through his connections, and he was 50 years old. It was a marriage of avarice. My uncle got a bribe of 200,000 rials, and my father got 160,000 in money and 300,000 in gold (dowry).…On the day of my marriage, I still had not reached puberty, and I reached puberty only in his house. He had intercourse with me on the first night, and of course, I did not know what a man does to a woman or how to act…my feelings on that night were horrible. I thought he was going to kill me, and he took me by force. I tried to defend myself that night, and I could not and then I fainted and lost consciousness and recovered only in the morning, and he left me alone to get rest.” (p. 462-463, [21]).

#### Tourist marriage

Tourist marriage, a recent manifestation of child marriage, involves a temporary union between a Yemeni girl and a man from a wealthy Gulf country. This phenomenon has arisen in response to the ongoing humanitarian crisis in Yemen stemming from the 2014 war. Tourist marriages can last from one day to several months and represent a temporary agreement rather than a legal marriage [7,21]. In a qualitative study involving stakeholders at NGOs focused on working and helping Yemeni women and children in crisis, a participant highlighted poverty as the primary factor contributing to the increasing incidence of this emerging form of child marriage [7].

> “Poverty, economic hardship, unemployment and increasing cost of living were all raised as reasons for families to resort to child marriage as a coping mechanism to permit them to alleviate poverty or the burdens of a large family with many daughters. Girls from low-income families are more likely to marry before 18” (p. 9, [7]).

#### Female genital mutilation and cutting

Five cross-sectional studies measured FGM/C, defined as ‘’all procedures involving partial or total removal of the external female genitalia or other intentional injuries to the female genital organs for non-medical reasons’’ [16,36,43,44,46]. Three studies assessed both women’s and their husbands’ attitudes toward FGM/C [16,44,46]. Estimates of the prevalence of FGM/C ranged from 16% in a 2008 subnational survey [44] and the 2013 DHS [12] to around 20% in the 1997 DHS [39] and the 2003 Arab Family Health Survey [43] and 89% in a population-representative FGM/C-specific survey in three southern Yemeni governorates [46]. Traditional birth attendants called *daya, gedah* , *kharshofa, or qabila* conducted FGM/C at home or in the community [16,44,46]. While some studies found maternal education and knowledge of the harms of FGM/C was predictive of their daughter’s likelihood of experiencing FGM/C [16,44], other studies did not [46].

#### Gender preference

The 1991/92 and 1997 DHS found that 20% and 33% of ever-married women preferred having a male over a female child [38,39]. In one qualitative study where 220 women aged <25 to >35 in Aden, Lahij, Hadramaut, Taiz, and Al Hudaydah governorates were asked about their gender preferences, respondents reported that they “pray to God” for a boy and not a girl child to make their husband happy [34].

> “I had nine daughters; my husband wants a son. I was so frightened, it’s a girl that I started a fever and was shivering from fright.” (pregnant woman, aged 33, urban residence) (p. 7, [34])
>
> “My husband is only waiting to hear the good news that the baby is a boy; he never thinks of me, only the gender of the baby, and I feel very badly.” (woman with three living children, aged 22, rural residence) (p. 7, [34])

#### Gender inequitable access to food, medical care, and education

Six studies described gender inequities in access to food, medical services, or education using data from qualitative studies [42,52–54], the DHS [35], the Pan Arab Project for Family Health (PAPFAM) [42], and the 2016 Household Income, Expenditure and Consumption Surveys (HIECS) [45]. A cross-sectional quantitative study on gendered access to education with 8–15-year-olds in 20 Yemeni districts found that over 30% of girls were not able or allowed to attend school, compared to 10% of boys [42]. A cross-sectional, multi-country survey on gendered access to medical care, found that Yemeni boys exhibited a higher likelihood of seeking or receiving hospital care compared to girls (prevalence sex ratio 0.85; p<0.05) [35]. A statistically significant proportion of women (3.66 on a scale of 1 to 5; p<0.001) reported feeling guilty for accessing medical services and were less likely than their male counterparts to spend money on medical care [53]. A 2005 cross-sectional study on gender and literacy as determinants of care-seeking behavior found that 63% of low-literacy women delayed diagnosis of tuberculosis (TB) (longer than eight weeks) compared to 44% of fully literate women [52]. In a 2007 study, most women (69%) reported having been harmed by family members when seeking medical care within the formal health system [53]. The two cross-sectional surveys mentioned above also found that women must seek permission from their husbands or male family members before visiting a doctor or clinic [52,53].

#### Child abuse or maltreatment

Seven studies measured violence against children (VAC) at home or in school, including emotional, physical, and sexual abuse [33,40,47–51]. The majority of the studies were cross-sectional. Four studies conducted interviews with children to gather information about their own experiences of abuse [21,47,48,51]. Two studies analyzed caregiver (mothers, aunts, grandmothers, guardians, teachers) reports of child abuse, either abuse that they had witnessed or been told about [40,50]. One study examined MICS data to determine the different forms of child abuse and related risk factors [40]. A high level (around 80%) of adults reported experiencing emotional abuse in their childhood across studies [40,47,49–51]. The 2013 DHS reported that 42% of children aged 2-14 (n=5,731) were exposed to severe physical punishment (being hit on the head or face with hands or by an implement) in the month before the survey. Nearly 80% (n=10,774) faced some form of physical discipline in the same timeframe [12]. The 2006 MICS reported that 94% of children aged 2-14 (n=2,702) were ever exposed to at least one form of physical or psychological abuse, and over 20% of children aged 5-14 (n=1,733) were engaged in child labor [41]. Three studies found a high prevalence of emotional and physical abuse among school children in Sana’a and four randomly selected districts of Aden [47,48,51]. A 2011 study of the different forms of child abuse prevalent in Yemen found that more than 90% of children aged 2-14 ever experienced emotional abuse [40], and a 2019 cross-sectional study found that 35% of students aged 18-24 reported experience of sexual abuse during their childhood [49]. A 2008 study of a random sample of students aged 7-10 (n=1,196) and another 2019 study with a random sample of students from five colleges at Hadramaut University (n=395) found shouting, yelling, hitting by hands, severe physical punishments, and sexual abuse to be commonly experienced by children [49,50]. Close to 60% of the interviewed mothers (n=119) of school-aged children living in rural areas reported hitting their children with their hands to discipline them [50]. Thirty-five per cent of students in one quantitative study (n=139) experienced sexual violence during childhood, defined as being “spoken to in a sexual way, unwanted sexual touching, forced to watch movies, take naked photos” [49]. Eighty per cent of students experienced verbal sexual harassment (n=111), and 19% (n=26) were coerced into watching pornographic movies [49].

### Perpetrators of violence

Most studies reported child abuse perpetrators as parents or other family members, teachers, and school principals in schools or universities [40,47–51]. FGM/C practices were mainly carried out by family, traditional healers, and community members at home or in health facilities [16,36,43,44]. Both partners and parents perpetrated honor violence [32].

### Legal context

Legal protections for women and children facing violence are limited. In cases of sexual violence against women or girls, families might force their daughters to marry the perpetrator as a means of restoring the family’s honor [7,21]. Divorced women are often blamed for their divorce [32]. One 2002 qualitative study found that Yemeni women felt that:

> ’Women have no rights…We are second-class citizens. Nobody listens, nobody cares, people think this is normal, but it is not normal" (p.343, [32]).

In the same study, a participant reported,

> “’I do not believe what I hear from the media about equality between man and woman in our society. Our society is a men’s society. Men decide what is right and what is wrong, not only in the family but in the whole society. They interpret the holy Qur’an in harmony with their interest. We know that our religion (Islam) guarantees many rights for women, but men accept only what is good for them and do not accept what is not good for them. For example, in some parts of Yemen, the community does not accept women’s inheritance: this is against Islamic law! This is because they don’t accept women as people” (p.343, [32]).

While Yemen is considering a bill that specifies girls should be at least 18 to marry, the bill has not been passed into law. In addition to country-level legal prohibitions against child marriage, alternative approaches that leverage Islamic prohibitions against child marriage are needed [21]. According to a workshop conducted by the Arab Resource Collective (ARC), early marriages are practiced due to weak laws and the inability to feed children [21]. The lack of birth or marriage registries makes it challenging to track the girls’ age at marriage [7,21], and the ongoing conflict has set back previous progress toward reducing child marriage in Yemen.

> “A boy and a girl can be considered adults, making it difficult for them to understand that they are marrying children. We managed to include it in the national dialogue, and we succeeded. But unfortunately, [child marriage] is now on hold due to conflict” (p. 10, [7]).

In a study aiming to gather participant suggestions for mitigating child marriage, recommendations included providing financial support to families for girls’ education, funding NGOs dedicated to addressing child marriage, raising awareness within communities about girls’ rights, and implementing local interventions as potential strategies [7].

### Quality assessment

The quality of primary research studies, excluding population-representative studies like DHS and MICS, is summarized in Table S5. The quantitative studies on corporal punishment in schools and experience of child maltreatment and abuse followed similar well-described study designs and sampling frames [40,47–50], but were small in scale and not representative. Most of the included qualitative studies had clearly stated research questions, methods, and findings. However, one article did not specify how the analysis was coded [33], and neither the research question nor methods were clear in one article [31].

### Ethical conduct of VAWC research

We adapted the rubric developed by Peterman, et. al., [58] to assess studies’ compliance with established norms for the ethical conduct of VAWC research, described in Table 2. Ethical concerns for VAWC research from the DHS and MICS studies is described elsewhere [59–62]. Studies that asked children or young adults about their experience of violence at home (N=3; 12,46,48) or in the classroom (N=3; 44–46) reported ethical clearance, verbal assent from participants, and used paper-based forms that did not collect identifying information. In two studies, where the participants were minors, consent was obtained from the parents [47,48]. The two qualitative studies that interviewed women and girls about their own experience of violence, including sexual violence, did not mention ethical approval or informed consent [21,32]. One of the studies mentioned that interviewers were trained psychology students and that they were accompanied by other women for safety purposes [32]. One quantitative study that interviewed male and female heads of household about their attitudes towards FGM/C, and in the case of women, their own experience of FGM/C, mentioned interviewer training, but did not specify what the training consisted of [46]. One quantitative survey of childhood experience of violence reported that the study team was trained to recognize any signs of distress exhibited as students completed the questionnaire [51]. No study mentioned making safety planning, resource lists, or support services available to participants. In one 2015 study, where female participants were sometimes accompanied by their husbands or older relatives, a second round of interviews was conducted when privacy could be ensured [34]. In resource-limited settings discussing the policy relevance of the findings is especially important to ensure ethical conduct and impact of future research, but most studies did not address the policy implications of their work.

**Table 2.** Ethical conduct of primary VAWC studies.

| Author, year of publication | Population | Focus | Ethics, interviewer training, safety planning |
| --- | --- | --- | --- |
| Al Taj, 2023 [46] | Male and female heads of household | Prevalence and attitudes towards FGM/C | Ethical review by Sana'a University, multistage random sampling, verbal IC, mentions interviewer training, but not what interviewers were trained in, interviews conducted in subject's home without others present. |
| Bamatraf, 2019 [49] | University students | Prevalence and risk factors for | Ethical review by Hadramaut University School of Medicine, verbal IC, self-completed, anonymous paper |
|  |  | child abuse | survey using validated instrument, returned in ballot box for anonymity |
| Ba-Saddik, 2012 [47] | Secondary school students | Prevalence of and risk factors for emotional abuse in classrooms | Ethical review by Committee of Research and Postgraduate Studies, Faculty of Medicine and Health Sciences, Aden University, written IC from parents, verbal assent from participants, self-completed, anonymous paper survey using validated instrument. |
| Ba-Saddik, 2013 [48] | Secondary school students | Prevalence of and risk factors for physical abuse in classrooms | Ethical review by Committee of Research and Postgraduate Studies, Faculty of Medicine and Health Sciences, Aden University, Written IC from parents, verbal assent from participants, self-completed, anonymous paper survey using validated instrument. |
| Ba-Obaid, 2002 [32] | Women aged 15-55 | Types of VAW | Multistage sample, trained female interviewers who were psychology students, sometimes accompanied by a second interviewer for safety reasons, interviews conducted in respondent home when adult males are not present. |
| Alizzy, 2017 [51] | Students aged 11-16 | Relation between family violence and child behavioral issues | Ethical review from Faculty of Arts and Humanities at Sana'a University, verbal assent from participants, self-completed, anonymous paper survey using validated instrument, participants debriefed after completion of the questionnaire, data collection team trained to handle signs of distress during survey completion. |
| Ouis, 2009 [21] | Adolescents aged 12-18 | Experience of and attitudes towards honor violence, sexual abuse, early marriage | Convenience sample of participants who had experienced sexual violence, no mention of IC, ethical clearance, resources or support, etc. |

IC=Informed Consent

#### Barriers, tools, strategies, and resources for addressing and reporting violence, including safety planning

One 2002 study found that only 3% of women who experienced violence went to the police, preferring to reach out to parents or relatives or not seek any external support rather than the police or legal support [32]. Two studies reported that women could only go to the police when accompanied by a male household member, leading to underreporting of GBV [32,51]. Most studies did not ask women or children about reporting or whether they accessed any resources to address the violence. Religious and cultural norms were identified as the most critical barriers to reporting most types of VAWC [16,36,43,44]. A cross-sectional study that asked dentists about their intention to report suspected VAWC found that only 60% of dentists would report VAWC to the police, followed by other agencies in Yemen [55]. A 2017 AMRO regional study by UNICEF reported that a shortage of skilled NGO personnel and a lack of awareness of available services were key barriers to accessing resources, as NGOs addressing gender-related issues do not have the staff to effectively manage reported GBV cases and do not well understand how to connect women to services or what services are available [7]. The same study found that girls facing GBV and child marriage experience limitations to their mobility due to gender norms and restricted access to money needed to pay for resources that are generally located outside of their community [7].

Sexual abuse is highly stigmatized in Yemen, which limits reporting. One study found that women or girls who were sexually abused face “severe punishment” from their parents, who might “kill them” or “force them to marry rapists” [21]. The same study found that adolescent girls in Yemen felt that women and girls could be considered responsible for the sexual violence they experienced [21]. Two studies highlighted that a lack of education for girls and women increases their exposure to violence from husbands compared to more educated women [32,33]. Women reported the widespread cultural beliefs that they are second-class citizens and belong to men, which likely complicates their ability to report violence [32]. Poor implementation of the FGM/C law in Yemen, lack of educational infrastructure, lack of employment opportunities, and the ongoing conflict were also cited as barriers to reporting [44]. Lack of awareness of FGM/C status, which is at a very young age, was also cited as a barrier to reporting [16]

### Discussion

This systematic review synthesizes evidence from qualitative and quantitative studies on the types, prevalence, perpetrators of, and risk factors for VAWC in Yemen. The 30 studies included in the analysis collected data on a range of forms of VAWC, including honor violence, female genital mutilation and cutting, early and very early marriage, tourist marriage, family and intimate partner violence, school-based violence, and gender inequities in access to food, education, and medical care. Included studies reported a high prevalence of many forms of violence, including corporal punishment in schools and intimate partner violence. We identified seven primary research studies that asked women or children about their experience of violence [21,32,46–49,51] and nine studies that reused data from the Yemen DHS, MICS, PAPFAM, and other national and sub-national household surveys [16,35–37,40,42–45]. Six additional studies asked Yemeni dentists about reporting violence [55], students about their attitudes towards women living with HIV [56], gender differences in the prevalence of depression [54], women’s perceptions of self-worth in relation to health-seeking behavior [53], tuberculosis patients on how gender and literacy affect healthcare access [52], and mothers on the factors that cause their fear of childbirth [34]. Studies measured IPV and non-partner family violence (both physical and psychological), sexual violence, early marriage, tourist marriage, child maltreatment, FGM/C, honor violence, child labor, school-based violence, including corporal punishment, and gender inequities in access to health care, education, and food. Studies had very different designs and sampling frameworks, preventing direct comparisons.

Similar to global trends [63], we found that IPV and other forms of family violence were the most prevalent forms of violence reported in quantitative studies of different types of VAWC. Studies on structural violence in Yemeni society [31,33] linked political instability and social exclusion to GBV, including higher rates of violence and social stigma directed towards marginalized ethnic groups, like the Muhamasheen.

#### Risk Factors

Risk factors for VAWC included gender inequitable norms and laws, economic instability, lower SES, rural residence, low levels of parental education, and low levels of education in women and girls. IPV and FGM/C were linked to a lack of maternal education, cultural norms and traditions in Yemeni society and, in the case of IPV, restrictive divorce laws. FGM/C was linked to mothers’ experience of FGM/C and early marriage. Child maltreatment and early child marriage were related to low levels of parental education and markers of low SES, including larger household size and reduced household income. These findings align with the determinants of VAWC in humanitarian crises in other countries, where cultural, economic, and educational factors are related to the incidence of GBV [64,65]. For example, studies in Sudan and Egypt found that lack of parental education, older age (where older women are more likely to have undergone FGM/C), and patriarchal norms were associated with FGM/C [66,67]. One study included in the review compared attitudes towards honor violence between Yemen, the occupied Palestinian Territories, and Lebanon and found that adolescent girls from Yemen and the Palestinian Territories were more likely than Lebanese girls to believe that women and girls were themselves responsible for their exposure to sexual violence [21].

### Barriers to reporting VAWC

Barriers to reporting violence included the lack of gender equitable laws, services or support for women or children who experience violence, and gender-inequitable child custody laws. Studies found that violence, when reported, was reported to family members or the local Sheikh, which is consistent with another study that found stigma and lack of support for women who experienced violence led to underreporting [68]. Yemen could consider female police officers to address that barrier to reporting violence [32]. The study also found that educational programs and providing mental health services were effective in reducing violence [68].

### Violence against3 children

Preschool & school-aged children and adolescents reported a higher prevalence of emotional rather than physical abuse, which is consistent with research conducted in South Africa [69,70]. Studies in this review reported low levels of sexual violence against children, which is mirrored in global research and likely related to underreporting [71–73]. Included studies reported that male children experienced higher levels of maltreatment than female children, which was similar to findings from another study conducted in Iran [74] but differed from findings in other Arab countries, including Tunisia and UAE [75]. Although child labor is a significant concern in Yemen, only the 2005 MICS measured this form of violence against children. Rates of corporal violence at school and child experience of maltreatment or physical abuse seemed much higher in Yemen than in related contexts, such as Sudan and Syria [76]. One study indicated that children in rural areas encountered elevated levels of corporal punishment in schools [50], which was supported in another study in Mali and Libya [77]. Risk factors associated with child abuse and maltreatment, including gender, older age, rural residence, family socioeconomic status, family size, parent’s education, and parent’s alcohol use, were similar to those reported in a study in Saudi Arabia [78] and to findings from conflict-affected countries, including Pakistan [79] and Myanmar [80].

#### Sexual violence

Sexual violence is a common yet highly stigmatized form of VAWC and is subject to high levels of underreporting [21]. Tourist marriage is a new form of GBV that is a last resort for families with limited resources to feed their families. Tourist marriage was also identified as an emerging concern in a study in Giza, Egypt [81]. Policymakers suggest that poverty reduction and community education [82] can help reduce early marriage in Yemen and other contexts in the Eastern Mediterranean, South and Southeast Asia and Africa [83].

#### Female genital mutilation and cutting

Yemen has made strides to prevent FGM/C; however, the government’s statement against FCM/C falls short of a nationwide law [44]. In contrast, Egypt has passed a law that prohibits FGM/C and punishes perpetrators [44,67]. Women’s empowerment, the involvement of religious leaders, and the education of parents and communities have been cited as pathways towards reducing FGM/C [16,36,43,44].

#### Gender inequities in access to medical care, education

Several included studies found gender inequities in healthcare access [35,52,53], education [45], and nutrition [37]. These studies are similar to those in other gender inequitable settings where women and girl children have lower levels of access to food [84], health care [85], and education [86] than males.

### Strengths & limitations

This is the first mixed methods systematic review to summarize findings on different forms of VAWC in Yemen. We searched four biomedical databases and NGO websites and used snowball sampling to identify primary and secondary research on VAWC. Limitations stem from the predominantly cross-sectional design of most studies, constraining the ability to infer causality. Future research should focus on longitudinal studies to better understand the dynamics of VAWC over time and explore the efficacy of various intervention strategies. The high levels of stigma surrounding VAWC, especially sexual violence, may have led to underreporting. Most primary research studies, other than the large population-representative surveys like DHS and MICS, did not report on interviewer training to ensure women and children could report on their experience of violence in a safe environment, free of stigma from the interviewer. Adequately funding studies and ensuring that well-funded studies have a coherent plan to address ethical issues in VAWC research could ensure studies on VAWC in Yemen uphold best practices in ethical research on VAWC in resource-limited contexts.

### Recommendations for future work

Most studies on VAWC were conducted before the 2014 war in Yemen. The war and pandemic have increased the frequency and forms of VAWC through multiple avenues. The humanitarian research and intervention communities should explore and address the causes and consequences of VAWC, and the international development community should consider investing in research to develop community-based efforts to support Yemeni women and children who experience or have experienced violence, including war-related violence and the associated trauma.

This review found significant ethical concerns related to existing research, including the absence of informed consent or ethical review. Only two studies, outside of the DHS and MICS, mentioned interviewer training, and none of the studies that interviewed women or children about their experience of violence discussed safety planning or connecting women and children to available resources. Given the lack of resources in Yemen, using positive deviance [87] approaches to identify strategies that would be possible and safe for women and children who experience violence to mitigate or prevent violence. Yemen is a complex context, and Yemeni women and children need to inform the design and execution of research and interventions.

### Policy implications

In keeping with policy-related research in Yemen [88], included studies identified severe poverty and displacement, cultural norms, societal stigma, and a lack of sufficient legal protections and support services as barriers to addressing VAWC in Yemen. Humanitarian support for Yemeni families in need is a crucial starting point to curb child and forced marriages [89]. Gender-inequitable child custody laws, cultural beliefs that normalize VAWC, and stigmatizing attitudes towards survivors of violence, including coercive marriages to preserve family honor in the context of sexual violence and blaming divorced women, [90] make addressing VAWC extremely complex. Addressing weak enforcement of protections for women and children [7] and gender inequitable laws or gaps in legislation, including no minimum age of marriage, should be part of a national strategy to address VAWC in Yemen [91]. A locally developed and supported policy action plan is essential for fostering lasting changes in gender-inequitable attitudes towards women and children [92]. This summary of the current landscape of VAWC-related research could support the design of a policy agenda that addresses barriers to measuring, reporting, and responding to VAWC. While not well described in the included studies, the large-scale conflict and related internal displacement have intensified GBV in Yemen [93]. Significant investment is needed in research and services, including basic needs, education, psychological services, and VAWC-related prevention and mitigation to support Yemeni women and children better. Similarly, child labour, an important form of violence that was not well explored in the included studies, should be considered in the research and development agenda [94]. By combining efforts to address gender inequitable laws and harmful cultural practices and providing holistic support to Yemeni families, Yemen can make strides towards reducing VAWC and creating a safer environment for women and children [95].

## Conclusions

VAWC is a global public health and human rights issue that affects the global community’s ability to achieve most of the 17 SDGs. Violence increases in unstable environments. Understanding and addressing VAWC is context-dependent, requiring meaningful community engagement, cross-sectoral support, and policy action. This SR highlights the high levels of VAWC in Yemen, driven by poverty, displacement, and gender inequitable norms and policies. While we know that VAWC increases during conflicts and when people are displaced, we did not identify any studies on VAWC that collected data after the onset of the ongoing conflict in Yemen, which represents a missed opportunity to understand how the ongoing war has reversed prior gains in reducing the prevalence of child and very early child marriage and introduced new forms of gender-based violence, including tourist marriage. The lack of funding for primary research on VAWC may be one reason why included studies did not report on interviewer training, mental health support for research participants, or safety planning, all of which should be included in VAWC-related research in humanitarian settings or fragile states. Addressing VAWC in Yemen will require a commitment on the part of the international community to fund locally-led efforts to determine immediate and long-term steps to alleviate the factors that drive VAWC (e.g., war, poverty), prevent and mitigate VAWC, and address the effects of VAWC on maternal and child health and related SDGs.

## Supporting information

S1 Text. Data extraction form

S2 Table. Database-specific searches

S3 Table. Articles excluded at full text stage

S4 Table. Overview of quantitative studies

S5. Table. Quality assessment

S1 Table. PRISMA Checklist

## Data Availability

All data generated in this systematic review are available in the article or its appendices.

## Acknowledgements

We want to thank the participants in the primary research studies included in this review for their courage and honesty in confronting the global challenge of VAWC and the NGOs and researchers working to ensure that their stories are heard and their needs addressed.

## Supporting Information

S1 Table. PRISMA checklist

S2 Table. Database-specific search strategies

S3 Table. List of articles excluded at full-text review stage with reason for exclusion S1 Text. Data extraction form

S4 Table. Overview of quantitative studies and quantitative section of mixed methods studies

S5 Table. Quality assessment of included studies

## Notes

### Competing Interest Statement

The authors have declared no competing interest.

### Clinical Protocols

https://www.crd.york.ac.uk/prospero/display_record.php?ID=CRD42021237855

### Funding Statement

MAZ received funding support from the Else Kroener-Fresenius-Stiftung within the Heidelberg Institute for Global Health at Universitaetsklinikum Heidelberg for one year of her work on this project (Award Number D10053008). The Heidelberg University Library supported the open-access fee for this article. The Else Kroener-Fresenius-Stiftung had no role in the study's design, conduct or reporting.

