## Supplementary material for "Violence against women and children in Yemen: A mixed-methods systematic review": S1 Text. Data extraction form

1) General study information
a) Author
b) Title (primary research study)
c) Publication year
d) Research question of primary research
e) Main purpose/aim/objective/motivation of study
2) Population
a) Location (city)
b) How was the population captured (schools, displacement camps, shelters, health care facilities/others)?
c) Governates/Districts/Institution/Department
d) Rural/urban setting in Yemen/not specified.
e) sample size/method of sampling/age/inclusion & exclusion criteria/study design/study type
3) Intervention or comparison groups
f) Intervention (if any)/comparison/outcome/key findings
g) operational definition of violence used in citations?
4) Outcomes measures
a) Type of violence and abuse/place of violence occurred/ perpetrators/ resources used to address/whom did they report/tools and strategies used to address/reasons or predictors of violence/risk factors/ subgroup affected/severity level of violence/ physical, psychological impact on survivors/violence assessment/statistical analysis/
b) Effect and occurrence measures (odds ratio, prevalence, incidence, risk ratio, etc.)/ statistical analysis/proportion of missing data/ how missing data dealt with (imputation or listwise deletion).
5) Ethics and confidentiality measures
a) Ethics approval/mode of consent/consent person/interview conducted person/strategies used for informed consent/prior logistic planning/interview responded person/composition of research teams/participant awareness level/language of instructions/status of reviewer/pilot study/examine of reliability and validity of survey.
6) Safety planning
   a) Security steps by research teams/ survey environment /trainings of research teams/ response towards women and children distress level/agreement in research teams and potential providers/use of loaded terms/confidentiality of data/concealing of identity/ethical obligations/safety approach on phases of implementation/emergency security plan/compensation received by participants?
7) Challenges and evidence
a) Attitudes, opinions of women and children towards violence/Cultural social, legal, political barriers observed/Potential risks and benefits/consequences of violence/gaps in evidence/implication of study/recommendations by researchers/ impact of research on intended beneficiaries/ limitation of study.
