## Supplementary material for "Violence against women and children in Yemen: A mixed-methods systematic review": S2 Table. Database-specific searches

### S2 Table. Database specific search strategy

### Ovid(Medline)

|  | **Mesh Terms** |
| --- | --- |
| 1 | Adolescent/ |
| 2 | Infant/ |
| 3 | Child/ |
| 4 | exp Women/ |
| 5 | Female/ |
| 6 | (child* or infant* or woman or women or female$ or adolescent* or girl* or teen$ or preteen or young people or youth or young person$ or juvenile$).ti,ab. |
| 7 | Battered women/ or Abused women/ |
| 8 | exp Domestic violence/ |
| 9 | exp Child Abuse/ |
| 10 | incest/ or rape/ or torture/ |
| 11 | Adverse Childhood Experiences/ |
| 12 | Gender-Based Violence/ |
| 13 | Gun Violence/ |
| 14 | Social Discrimination/ or Sexism/ |
| 15 | exp Intimate Partner Violence/ or spouse abuse/ or dating violence/ |
| 16 | Sexual Harassment/ or Physical abuse/ |
| 17 | Workplace violence/ |
| 18 | exp Sex offenses/ or exp homicide/ |
| 19 | Circumcision, Female/ |
| 20 | exp War Crimes/ |
| 21 | pandemics/ or coronavirus infection |
|  | **Text Terms** |
| 22 | ((batter$ or abuse*) adj3 (wom*n or wife or wives)).ti,ab. |
| 23 | ((domestic or home) adj2 violen$).ti,ab. |
| 24 | ((infan$ or child$ or teen$ or adolesc$ or minor$ or toddler$ or baby or babies) adj3 abuse* adj3 (maltreat$ or neglect$ or abuse* or physical* or sexual* or emotion*)).ti,ab. |
| 25 | (rape$ or torture* or incest*).ti,ab. |
| 26 | adverse child* experiences*.ti,ab. |
| 27 | ((gender or gender based* or peer) adj3 violen$).ti,ab |
| 28 | (gun violen* or dowry death$).ti,ab. |
| 29 | (sex$ or sexism or social discrimination*).ti,ab. |
| 30 | ((wife or intimate partner* or wives or partner* or spouse*or date or dating) adj3 (abuse$ or violen*)).ti,ab. |
| 31 | ((sex$ or physical) adj3 (harrasment$ or abuse* or violen*)).ti,ab. |
| 32 | ((workplace or social or community) adj3 violen*).ti,ab. |
| 33 | (sex offens* or homicid$ or femicide$ or murde$ or coerc$).ti,ab. |
| 34 | (female genital mutilation$ or female circumcision or female genital cutting or infibulation$).ti,ab. |
| 35 | (war violence* or war crime*).ti,ab. |
| 36 | (pandemics or coronavir* or COVID* or COVID 19* or coronovirus* or HCoV* or betacoronavir* or sars-cov* or sarscov* or sars-coronovirus* or "beta-coronavirus" or "beta-coronaviruses" or "corona virus" or "virus corona" or "corono virus" or "virus corono" or covid* or "2019-ncov" or cv19* or "cv-19" or "cv 19" or "n-cov" or ncov*).ti,ab. |
| 37 | Yemen/ |
| 38 | Yemen.ti,ab. |
| 39 | or/1-6 |
| 40 | or/7-36 |
| 41 | 37 or 38 |
| 42 | 39 AND 40 AND 41 |
| 43 | exp animals/ not humans/ |
| 44 | 42 not 43 |

### Ovid (Embase)

| SR. | MESH TERMS |
| --- | --- |
| 1 | Adolescent/ |
| 2 | Infant/ |
| 3 | Child/ |
| 4 | exp Women/ |
| 5 | Female/ |
| 6 | (child* or infant* or woman or women or female$ or adolescent* or girl* or teen$ or preteen or young people or youth or young person$ or juvenile$).ti,ab. |
| 7 | Battered women/ or Abused women/ |
| 8 | exp Domestic violence/ |
| 9 | exp Child Abuse/ |
| 10 | incest/ or rape/ or torture/ |
| 11 | 2 Adverse Childhood Experiences/ |
| 12 | Gender-Based Violence/ |
| 13 | Gun Violence/ |
| 14 | Social Discrimination/ or Sexism/ |
| 15 | exp Intimate Partner Violence/ or spouse abuse/ or dating violence/ |
| 16 | Sexual Harassment/ or Physical abuse |
| 17 | Workplace violence/ |
| 18 | exp Sex offenses/ or exp homicide/ |
| 19 | Circumcision, Female/ |
| 20 | exp War Crimes/ |
| 21 | pandemics/ or coronavirus infection |
|  | TEXT Terms |
| 22 | ((batter$ or abuse*) adj3 (wom*n or wife or wives)).ti,ab. |
| 23 | ((domestic or home) adj2 violen$).ti,ab. |
| 24 | ((infan$ or child$ or teen$ or adolesc$ or minor$ or toddler$ or baby or babies) adj3 abuse* adj3 (maltreat$ or neglect$ or abuse* or physical* or sexual* or emotion*)).ti,ab. |
| 25 | (rape$ or torture* or incest*).ti,ab. |
| 26 | adverse child* experiences*.ti,ab. |
| 27 | ((gender or gender based* or peer) adj3 violen$).ti,ab |
| 28 | (gun violen* or dowry death$).ti,ab. |
| 29 | (sex$ or sexism or social discrimination*).ti,ab. |
| 30 | ((wife or intimate partner* or wives or partner* or spouse*or date or dating) adj3 (abuse$ or violen*)).ti,ab . |
| 31 | ((sex$ or physical) adj3 (harrasment$ or abuse* or violen*)).ti,ab. |
| 32 | ((workplace or social or community) adj3 violen*).ti,ab. |
| 33 | (sex offens* or homicid$ or femicide$ or murde$ or coerc$).ti,ab. |
| 34 | (female genital mutilation$ or female circumcision or female genital cutting or infibulation$).ti,ab. |
| 35 | ( war violence* or war crime*).ti,ab. |
| 36 | (pandemics or coronavir* or COVID* or COVID 19* or coronovirus* or HCoV* or betacoronavir* or sars-cov* or sarscov* or sars-coronovirus* or "beta-coronavirus" or "beta-coronaviruses" or "corona virus" or "virus corona" or "corono virus" or "virus corono" or covid* or "2019-ncov" or cv19* or "cv-19" or "cv 19" or "n-cov" or ncov*).ti,ab. |
| 37 | Yemen/ |
| 38 | Yemen.ti,ab. |
| 39 | or/1-6 |
| 40 | or/7-36 |
| 41 | 37 or 38 |
| 42 | 39 AND 40 AND 41 |
| 43 | exp animals/ not humans/ |
| 44 | 42 not 43 |

### CINAHL (Cumulative Index to Nursing and Allied Health Literature)

| S1 | MH Adolescent OR Infant |
| --- | --- |
| S2 | MH Infant/ |
| S3 | MH Child/ |
| S4 | MH Women/ |
| S5 | MH Female/ |
| S6 | TI ((child* or infant* or woman or women or female* or adolescent* or girl* or teen* or preteen or young people or youth or young person* or juvenile*)) OR AB ( (child* or infant* or woman or women or female* or adolescent* or girl* or teen* or preteen or young people or youth or young person* or juvenile*)) |
| S7 | MH Battered women/ or MH Abused women/ |
| S8 | MH Domestic violence/ |
| S9 | MH Child Abuse/ |
| S10 | MH incest/ or rape/ or torture/ |
| S11 | MH Adverse Childhood Experiences/ |
| S12 | MH Gender-Based Violence/ |
| S13 | MH Gun Violence/ |
| S14 | MH Social Discrimination/ or Sexism/ |
| S15 | MH Intimate Partner Violence/ or spouse abuse/ or dating violence/ |
| S16 | MH Sexual Harassment/ or Physical abuse/ |
| S17 | MH Workplace violence/ |
| S18 | MH Sex offenses/ or homicide/ |
| S19 | MH Circumcision, Female/ |
| S20 | MH War Crimes/ |
| S21 | MH pandemic or covid-19 or coronavirus |
| S22 | TI ( ((batter* or abuse*) N3 (wom*n or wife or wives)) ) OR AB ( ((batter* or abuse*) N3 (wom*n or wife or wives)) ) |
| S23 | TI ( ((domestic or home) N2 violen*) ) OR AB ( ((domestic or home) N2 violen*) ) |
| S24 | TI ( ((infan* or child* or teen* or adolesc* or minor* or toddler* or baby or babies) N3 abuse* N3 (maltreat* or neglect* or abuse* or physical* or sexual* or emotion*)) ) OR AB ( ((infan* or child* or teen* or adolesc* or minor* or toddler* or baby or babies) N3 abuse* N3 (maltreat* or neglect* or abuse* or physical* or sexual* or emotion*)) ) |
| S25 | TI ( (rape* or torture* or incest*) ) OR AB ( (rape* or torture* or incest*) ) |
| S26 | TI adverse child* experiences* OR AB adverse child* experiences* |
| S27 | TI ( ((gender or gender based* or peer) N3 violen*) ) OR AB ( ((gender or gender based* or peer) N3 violen*) ) |
| S28 | TI ( (gun violen* or dowry death*) ) OR AB ( (gun violen* or dowry death*) ) |
| S29 | TI ( (sex* or sexism or social discrimination*) ) OR AB ( (sex* or sexism or social discrimination*) ) |
| S30 | TI ( ((wife or intimate partner* or wives or partner* or spouse*or date or dating) N3 (abuse* or violen*)) ) OR AB ( ((wife or intimate partner* or wives or partner* or spouse*or date or dating) N3 (abuse* or violen*)) ) |
| S31 | TI ( ((sex* or physical) N3 (harrasment* or abuse* or violen*)) ) OR AB ( ((sex* or physical) N3 (harrasment* or abuse* or violen*)) ) |
| S32 | TI ( ((workplace or social or community) N3 violen*) ) OR AB ( ((workplace or social or community) N3 violen*) ) |
| S33 | TI ( (sex offens* or homicid* or femicide* or murde* or coerc*) ) OR AB ( (sex offens* or homicid* or femicide* or murde* or coerc*) ) |
| S34 | TI ( (female genital mutilation* or female circumcision or female genital cutting or infibulation*) ) OR AB ( (female genital mutilation* or female circumcision or female genital cutting or infibulation*) ) |
| S35 | TI ( (war violence* or war crime*) ) OR AB (war violence* or war crime*) ) |
| S36 | TI ( (pandemics or coronavir* or COVID* or COVID 19* or coronovirus* or HCoV* or betacoronavir* or sars-cov* or sarscov* or sars-coronovirus* or "beta-coronavirus" or "beta-coronaviruses" or "corona virus" or "virus corona" or "corono virus" or "virus corono" or covid* or "2019-ncov" or cv19* or "cv-19" or "cv 19" or "n-cov" or ncov*) ) OR AB ( (pandemics or coronavir* or COVID* or COVID 19* or coronovirus* or HCoV* or betacoronavir* or sars-cov* or sarscov* or sars-coronovirus* or "beta-coronavirus" or "beta-coronaviruses" or "corona virus" or "virus corona" or "corono virus" or "virus corono" or covid* or "2019-ncov" or cv19* or "cv-19" or "cv 19" or "n-cov" or ncov*) ) |
| S37 | TI Yemen OR AB Yemen |
| S38 | MH Yemen |
| S39 | S1 OR S2 OR S3 OR S4 OR S5 OR S6 |
| S40 | S7 OR S8 OR S9 OR S10 OR S11 OR S12 OR S13 OR S14 OR S15 OR S16 OR S17 OR S18 OR S19 OR S20 OR S21 OR S22 OR S23 OR S24 OR S25 OR S26 OR S27 OR S28 OR S29 OR S30 OR S31 OR S32 OR S33 OR S34 OR S35 OR S36 |
| S41 | S37 OR S38 |
| S42 | S39 AND S40 AND S41 |
| S43 | TI ( ((animal* OR canine* OR dog* or feline* OR hamster* OR lamb* OR mice OR mouse OR monkey* OR murine OR pig* OR piglet* OR porcine OR primate* OR rabbit* OR rat* OR rodent* OR sheep* OR frog* OR worm* OR trematode) ) NOT TI ( (human* OR patient*)) ) |
| S44 | S42 NOT S43 |

### Web of Science (WoS)

| #1 | TS=Adolescent |
| --- | --- |
| #2 | TS=Infant |
| #3 | TS=Child |
| #4 | TS=Women |
| #5 | TS=Female |
| #6 | TI=(child* or infant* or woman or women or female* or adolescent* or girl* or teen* or preteen or young people or youth or young person* or juvenile*) OR Ab=(child* or infant* or woman or women or female* or adolescent* or girl* or teen* or preteen or young people or youth or young person* or juvenile*) |
| #7 | TS=(Battered women/ or Abused women/) |
| #8 | TS=Domestic Violence/ |
| #9 | TS=Child Abuse/ |
| #10 | TS=(incest/ or rape/ or torture/) |
| #11 | TS= (Adverse Childhood Experiences) |
| #12 | TS=(Gender Based Violence) |
| #13 | TS=Gun Violence |
| #14 | TS=(Social Discrimination/ or Sexism/) |
| #15 | TS=( Intimate Partner Violence/ or spouse abuse/ or dating violence/) |
| #16 | TS=(Sexual Harassment/ or Physical abuse/) |
| #17 | TS=(Workplace violence/) |
| #18 | TS=(Sex offenses/ or homicide/) |
| #19 | TS=(Circumcision, Female/) |
| #20 | TS=(War Crimes/) |
| #21 | TS=(pandemic or covid-19 or coronavirus) |
| #22 | TI=((batter* or abuse*) NEAR/3 (wom*n or wife or wives) ) OR AB=((batter* or abuse*) NEAR/3 (wom*n or wife or wives) ) |
| #23 | TI=((domestic or home) NEAR/2 violen*) OR AB=((domestic or home) NEAR/2 violen*) |
| #24 | TI=((infan* or child* or teen* or adolesc* or minor* or toddler* or baby or babies) NEAR/3 abuse* NEAR/3 (maltreat* or neglect* or abuse* or physical* or sexual* or emotion*) ) OR AB=((infan* or child* or teen* or adolesc* or minor* or toddler* or baby or babies) NEAR/3 abuse* NEAR/3 (maltreat* or neglect* or abuse* or physical* or sexual* or emotion*) ) |
| #25 | TI=(rape$ or torture* or incest*) OR AB=(rape$ or torture* or incest*) |
| #26 | TI=(adverse child* experiences*) OR AB=(adverse child* experiences*) |
| #27 | TI=((gender or gender based* or peer) NEAR violen*) OR AB=((gender or gender based* or peer) NEAR violen*) |
| #28 | TI=(gun violen* or dowry death*) OR AB=(gun violen* or dowry death*) |
| #29 | TI=(sex* or sexism or social discrimination*) OR AB=(sex* or sexism or social discrimination*) |
| #30 | TI=(wife or intimate partner* or wives or partner* or spouse*or date or dating NEAR/3 abuse* or violen*) OR AB=(wife or intimate partner* or wives or partner* or spouse*or date or dating NEAR/3 abuse* or violen*) |
| #31 | TI=((sex* NEAR harassment*) or (sex* NEAR abuse*) or (sex* NEAR violence) or (physical NEAR harassment) or (physical NEAR abuse) or (physical NEAR violence)) OR AB=((sex* NEAR harassment*) or (sex* NEAR abuse*) or (sex* NEAR violence) or (physical NEAR harassment) or (physical NEAR abuse) or (physical NEAR violence)) |
| #32 | TI=((workplace NEAR violen*) OR (social NEAR violen*) or (community NEAR violen*)) OR AB=((workplace NEAR violen*) OR (social NEAR violen*) or (community NEAR violen*)) |
| #33 | TI=(sex offens* or homicid* or femicide* or murde* or coerce*) OR AB=(sex offense* or homicid* or femicide* or murde* or coerc*) |
| #34 | TI=(female genital mutilation* or female circumcision or female genital cutting or infibulation*) OR AB=(female genital mutilation* or female circumcision or female genital cutting or infibulation*) |
| #35 | TI=(war violence* or war crime*) OR AB=( war violence* or war crime*) |
| #36 | TI=(pandemics* or coronavir* or COVID* or COVID19* or coronovirus* or HCoV* or betacoronavir* or sars-cov* or sarscov* or sars-coronovirus* or beta-coronavirus* or beta-coronaviruses* or corona virus* or virus corona* or corono virus* or virus corono* or covid* or 2019-ncov* or cv19* or cv-19* or cv19* or n-cov* or ncov*) OR AB=(pandemics* or coronavir* or COVID* or COVID19* or coronovirus* or HCoV* or betacoronavir* or sars-cov* or sarscov* or sars-coronovirus* or beta-coronavirus* or beta-coronaviruses* or corona virus* or virus corona* or corono virus* or virus corono* or covid* or 2019-ncov* or cv19* or cv-19* or cv19* or n-cov* or ncov*) |
| #37 | TS=Yemen/ |
| #38 | TI=Yemen OR AB= Yemen |
| #39 | #6 OR #5 OR #4 OR #3 OR #2 OR #1 |
| #40 | #36 OR #35 OR #34 OR #33 OR #32 OR #31 OR #30 OR #29 OR #28 OR #27 OR #26 OR #25 OR #24 OR #23 OR #22 OR #21 OR #20 OR #19 OR #18 OR #17 OR #16 OR #15 OR #14 OR #13 OR #12 OR #11 OR #10 OR #9 OR #8 OR #7 |
| #41 | #38 OR #37 |
| #42 | #41 AND #40 AND #39 |
| #43 | TS=(animal model OR animal* NOT human) |
| #44 | #42 NOT #43 |

TS=(Adolescent OR Infant OR Child OR women OR Female)

TI=(child* or infant* or woman or women or female* or adolescent* or girl* or teen* or preteen or young people or youth or young person* or juvenile*)

Ab=(child* or infant* or woman or women or female* or adolescent* or girl* or teen* or preteen or young people or youth or young person* or juvenile*)

TS=(Battered Women OR abused Women or Domestic Violence OR Child Abuse OR incest OR rape OR torture or Adverse Child Experiences OR gender Based violence or Gun Violence OR social discrimination or sexism or intimate partner violence or dating violence or sexual harassment or physical abuse or workplace violence or sex offenses or homicide or female circumcision or war crimes or pandemic or covid-19 or coronavirus)

TI=(wife or intimate partner* or wives or partner* or spouse*or date or dating NEAR/3 abuse* or violen*)

AB=(wife or intimate partner* or wives or partner* or spouse*or date or dating NEAR/3 abuse* or violen*)

TI=((sex* or physical) NEAR (harassment* or abuse* or violen*))

AB=((sex* or physical) NEAR (harassment* or abuse* or violen*))
