## Supplementary material for "Violence against women and children in Yemen: A mixed-methods systematic review": S3 Table. Articles excluded at full text stage

### S3 Table. List of articles excluded at full text review stage with reason for exclusion

| **Title of studies excluded** | **Reason for exclusion** |
| --- | --- |
| Child protection assessment in humanitarian emergencies: Case studies from Georgia, Gaza, Haiti and Yemen | Systematic review, commentary, editorial or other non-primary research |
| Child abuse and neglect in the Arab Peninsula | Systematic review, commentary, editorial or other non-primary research |
| Between Worlds: A Yemeni Mother's Experiences of War, Separation, and Resettlement | Systematic review, commentary, editorial or other non-primary research |
| Increased Efforts by Modern States to Improve Their Reputations for Enforcing Women's Human Rights | Systematic review, commentary, editorial or other non-primary research |
| Expressed concerns of Yemeni adolescents | No measure of violence or stigma against Yemeni women and children |
| Female genital cutting. Evidence from the Demographic and Health Surveys | Systematic review, commentary, editorial or other non-primary research |
| A "synchronised attack' on life: the Saudi-led coalition's "hidden and holistic' genocide in Yemen and the shared responsibility of the US and UK | Systematic review, commentary, editorial or other non-primary research |
| Khat in East Africa: Taking women into or out of sex work? | No measure of violence or stigma against Yemeni women and children |
| In the Best Interests of the Child: Preventing Female Genital Cutting (FGC) | Systematic review, commentary, editorial or other non-primary research |
| WAYS TO COME, WAYS TO LEAVE Gender, Mobility, and Il/legality among Ethiopian Domestic Workers in Yemen | No measure of violence or stigma against Yemeni women and children |
| Deaths due to jambia-inflicted lesions in a domestic environment | Systematic review, commentary, editorial or other non-primary research |
| COVID-19 pandemic in Yemen: A questionnaire-based survey, what do we know? | No measure of violence or stigma against Yemeni women and children |
| Gender Differences in Worry About a Terrorist Attack: A Cross-National Examination of Individual- and National-Level Factors | No measure of violence or stigma against Yemeni women and children |
| Perspectives: Being a Female Dentist in Yemen | Systematic review, commentary, editorial or other non-primary research |
| Islamism, Secularism and the Woman Question in the Aftermath of the Arab Spring: Evidence from the Arab Barometer | No measure of violence or stigma against Yemeni women and children |
| For the sake of purity (and control). Female genital mutilation | Systematic review, commentary, editorial or other non-primary research |
| Women's rights, a tourist boom, and the power of khat in Yemen | Systematic review, commentary, editorial or other non-primary research |
| Gender Ideals in Turbulent Times: An Examination of Insecurity, Islam, and Muslim Men's Gender Attitudes during the Arab Spring | Systematic review, commentary, editorial or other non-primary research |
| Explaining divergent outcomes of the Arab Spring: the significance of gender and women's mobilizations | Systematic review, commentary, editorial or other non-primary research |
| The ongoing violence against women: Female Genital Mutilation/Cutting | Systematic review, commentary, editorial or other non-primary research |
| "The War Took Us Backwards" Yemeni Families and Dialectical Patriarchal Reordering | No measure of violence or stigma against Yemeni women and children |
| Women's health. The issue of unspoken abuse | Systematic review, commentary, editorial or other non-primary research |
| Defibulation During Vaginal Delivery for Women With Type III Female Genital Mutilation | No measure of violence or stigma against Yemeni women and children |
| Perinatal health in MENA area | Systematic review, commentary, editorial or other non-primary research |
| Legal protection for women in Yemen: A sorry state | Systematic review, commentary, editorial or other non-primary research |
| When Rebels Attack: Quantifying the Impacts of Capturing Territory from the Government in Yemen | No measure of violence or stigma against Yemeni women and children |
| Reproductive, maternal, newborn and child health service delivery during conflict in Yemen: a case study | No measure of violence or stigma against Yemeni women and children |
| Male Migration and 'Left-behind' Women: Bane or Boon? | No measure of violence or stigma against Yemeni women and children |
| 2017: a year in review | Systematic review, commentary, editorial or other non-primary research |
| Supporting female scientists in Yemen | Systematic review, commentary, editorial or other non-primary research |
| <Yemen 1996 MICS Summary (List of Indicators and Questionnaires)_English.pdf> | No measure of violence or stigma against Yemeni women and children |
| Early Marriage and Less Education as Independent Predictors for High Fertility in Yemen | No measure of violence or stigma against Yemeni women and children |
| Measuring Violence Against Women: A Global Index. | No measure of violence or stigma against Yemeni women and children |
