## Supplementary material for "Violence against women and children in Yemen: A mixed-methods systematic review": S4 Table. Overview of quantitative studies

**Table S4. Overview of quantitative studies and quantitative section of mixed-method studies**

| **Author, publication year** | **Violence (Scale used)** | **Violence subtypes** | **Prevalence/ per cent/mean of sample** | | | | **Uncertainty** | **Main findings** |
| --- | --- | --- | --- | --- | --- | --- | --- | --- |
|  |  |  | **Source, year** | **Age group** | **N** | **%** | **P value; 95% CI** |  |
| Akmatov, 2011 [1] | Child abuse (CTS-PC) | Psychological abuse | MICS 2006 | Mothers (aged 15-49) of children aged 2-14 | 2845 | 92% | NA | Male gender of child, urban residence, low SES and high number of household numbers associated with higher risk of all forms of child abuse |
|  |  | Moderate physical abuse |  |  |  | 81% |  |  |
|  |  | Severe physical abuse |  |  |  | 61% |  |  |
| Al Taj, 2023[2] | FGM/C | Women who have undergone FGM/C | 2020 survey | Male and female heads of household or oldest child if head of household absent (age not specified) | 646 women; 345 men | 89% | 84%-93% | Older maternal, own experience of FGM/C, positive, maternal or paternal attitude towards FGM/C, poor knowledge of harms of FGM/C, and rural residence associated with increased odds of youngest daughter having experienced FGM/C. |
|  |  | Daughter underwent FGM/C |  |  |  | 80% | 74%-85% |  |
| Alansari, 2006 [3] | Gender differences in mental health issues (BDI-II) | Physical and emotional abuse as factors | 2006 | University of Aden undergraduate students aged 18-25 | 648 | NA | p=0.52 | No significant associations between gender and depression |
| Alizzy, 2017 [4] | Family violence (NA) | Experiencing/witnessing physical abuse (Boys n=295, Girls n=303) | Interviews; 2013-2014 | Students aged 11-16 | 598 | Boys experiencing: 78%, witnessing: 32%;  Girls experiencing: 37%, witnessing: = 33% | Experiencing: p=0.0001  Witnessing: p=0.76 | Male gender related to higher probability of experiencing physical and emotional abuse, no association between gender and witnessing abuse. Girls exposed to abuse exhibited emotional difficulties (internalizing) while boys exhibited aggressive behaviors (externalizing). |
|  |  | Experiencing/witnessing psychological abuse (Boys n=295, Girls n=303) |  |  |  | Boys experiencing: 82%, witnessing: 79%  Girls experiencing: 61%, witnessing 79% | Experiencing: p=0.0001  Witnessing: p=0.91 |  |
|  |  | Low literacy |  |  |  | 31% |  |  |
| Al-Khulaidi, 2013 [5] | FGM/C (DHS) | Women who have undergone FGM/C | DHS 1997 | Women aged 15-49 who have heard of FGM/C | 5226 | 45% | p<0.001 | Maternal experience of FGM/C, low maternal literacy, and mother and father support for FGM/C positively associated and likelihood of daughter undergoing FDM/C. |
|  |  | Most-recently-born daughter underwent FGM/C |  |  |  | 29% |  |  |
|  |  | Women who have undergone FGM/C | FHS 2003 | Women aged 15-49 who have heard of FGM/C | 6302 | 38% | p<0.001 |  |
|  |  | Most-recently-born daughter underwent FGM/C |  |  |  | 22% |  |  |
| Alosaimi, 2019 [6] | FGM/C (HBMNC) | Women who have undergone FGM/C | HBMNC 2008–2009 | Women aged 15-49 who have heard of FGM/C | 4238 | 48% | p< 0.01 | Older maternal age, low maternal literacy and SES, and maternal early marriage, associated with higher odds of having undergone FGM/C and daughters being circumcised. Housing quality inversely related to likelihood of FGM/C. |
|  |  | Daughter underwent FGM/C |  |  |  | 34% |  |  |
| Alyahri, 2008 [7] | Child maltreatment (SDQ) | Shout or swear at children | 2002/2003 academic year | Parents and teachers of Yemeni school-aged children (7–10) | 1196 | Urban: 49%; Rural: 30% | p < 0.05;  p < 0.001 | Male gender, rural residence, lower maternal education and larger number of children in household related to higher probability of experiencing harsh physical punishment. |
|  |  | Hit child with hand/ hit child with implements |  |  |  | Urban: 52% Rural: 59% / Urban: 23%; Rural: 58% |  |  |
|  |  | Severe corporal punishment |  |  |  | Urban: 23%; Rural: 59% |  |  |
| Assaad, 2019 [8] | Gendered access to education (HIECS) | Female and male children who have ever attended school | 2006 from HIECS | Parents of Yemeni children aged 12-18 | 19168 | Female: 78%; Male: 97% | NA | Female children less likely than male children to attend or continue in school. |
|  |  | Female and male children who are currently attending school |  |  |  | Female: 44%; Male: 73% |  |  |
| Badahdah, 2016 [9] | HIV-related stigma (Yemeni AIDS Stigma Scale) | Attitudes towards people living with HIV | - | Undergraduate student with mean age of 21.52 | 613 | 80% believed WLWH should be sterilized; 62% believed that pregnant WLWH should be forced to get an abortion | NA | Male more likely than female students to stigmatize PLWH and to support restrictions on WLWH’s SRHR. |
| Bamatraf, 2019 [10] | Child abuse (ICAST) | Emotional abuse | 2015-2016 | Hadhramout University students aged 18-24 | 395 | 79% | NA | Male gender, witnessing violence at home, parents’ psychological issues positively correlated with experience physical, emotional, and sexual abuse. |
|  |  | Physical abuse |  |  |  | 76% |  |  |
|  |  | Sexual abuse |  |  |  | 35% |  |  |
| Ba-Obaid, 2002 [11] | DV; societal violence (Likert scale) | Threats | 1996-1997 | Yemeni women aged 15-55 | 111 | 51% | NA | Divorced or unemployed women more likely than married or employed women to experience physical violence. Husbands most common perpetrators of violence. |
|  |  | Sexual violence |  |  |  | 17% |  |  |
|  |  | Physical abuse |  |  |  | 55% |  |  |
|  |  | Restriction of freedom |  |  |  | 28% |  |  |
|  |  | Property damage and theft |  |  |  | 34% |  |  |
| Ba-saddik, 2012 [12] | Child abuse at schools (ICAST-CT) | Emotional abuse | 2009–2010 | Yemeni students aged 12-17 | 1066 | 55% | p<0.05/; 52.1 - 58.2 | Male gender, older age, lower level of education, father with lower level of education, higher number of household members associated with higher likelihood of experiencing violence. |
| Ba-Saddik, 2013 [13] | Child abuse at schools (ICAST-CT) | Physical abuse | 2009–2010 | Yemeni students aged 12-17 | 1066 | 56% | p<0.05 | Male gender, older age, lower level of education, father with lower level of education, higher number of household members associated with higher likelihood of experiencing violence. |
| Chamberlain, 2007 [14] | Gender inequity in healthcare access (NA) | Women experiencing guilt when money is expended on their health | 2007 | Yemeni women aged 15-45 | 100 | 3.66 | p<0.001; SD 1.3 | Female gender associated with lower levels of health care seeking. |
|  |  | Women who have been harmed by a family member |  |  |  | 69 | p < 0.001 |  |
|  |  | Ability of women to make their own decisions regarding health |  |  |  | 0 | p < 0.001 |  |
|  |  | Women with knowledge about their health rights |  |  |  | 40 | p < 0.001 |  |
| Costa, 2017 [15] | Gendered access to health care for children (DHS) | Gender inequity in care-seeking for children (sex ratio) | DHS 2013 | Preschool children’s | 15,383 | NS | 0.85 (0.78–0.92); p=0.000 | Female gender associated with lower levels of health care seeking. |
| Date, 2005 [16] | Gender inequity and illiteracy affecting healthcare access (NA) | Gender inequity in successful treatment outcomes | Interview; 2001- 2002 | PTB patients | 74 | Male: 46%; Female: 70% | p=0.046 | Male, low literacy patients less likely to complete treatment than female patients. |
|  |  | Gender and literacy inequity in diagnostic delay and unsuccessful treatment outcome (%) |  |  |  | Low literacy male: 55%; High literacy male: 17%; Low literacy female: 21%; High literacy female: 0% | p < 0.001 |  |
| El Tantawi, 2018 [17] | Intention to report suspected domestic/ physical violence (NA) | No subcategory | 2016 | Male and female Yemeni dentists, mean age of 31 | 291 | 60% | p < 0.0001 | Female gender, lack of perceived ability to identify violence survivors and lack of knowledge of mandatory reporting roles associated with lower perceived likelihood of reporting abuse. |
| Lewis, 2012 [18] | Youth violence and criminality, GBV (NA) | GBV | 2010-2012 | 18-49 yrs. | 61 | NA | NA | Victims of sexual violence face high levels of stigma leading to very low levels of reporting. |
| Liang, 2016 [19] | FGM/C (DHS) | Girls who have undergone FGM/C | DHS 2013 | Women aged 15-49 | 25,434 | 16% | NA | Maternal experience of FGM/C and child marriage, low maternal education, related to higher likelihood of daughter experiencing FGM/C |
| Sharaf, 2019 [20] | Gender inequity in child nutrition (DHS) | Mothers having a son preference | DHS 2013 | Mothers of children aged 0-5 years | 11,100; 12289 (for different regression analyses) | 22% | NA | No association |
| Smits, 2013 [21] | Gender inequity in education participation (NA) | Non-participation of girls (at age 11) | PAPFAM 2003 | Mothers (aged 16-49) of children aged 8-15 | NS | 30% | NA | Female children less likely to attend school in rural (44% vs 17%) and urban (9% vs 6%) |
|  |  | Non-participation of boys (at age 11) |  |  |  | 10% |  |  |
|  |  | Experiencing/witnessing psychological abuse (Boys n=295, Girls n=303) |  |  |  | Boys experiencing: 82%, witnessing: 79%  Girls experiencing: 61%, witnessing 79% | Experiencing: p=0.0001  Witnessing: p=0.91 |  |
|  |  | Low literacy |  |  |  | 31% |  |  |
| Yoder, 2013 [22] | FGM/C (FHS) | Women who have undergone FGM/C | FHS 2003 | Women aged 15-49 | 4,250,558 | 22% | NA | 931,899 Yemeni girls at risk of FGM/C |

NA=not applicable; MICS=Multi-Indicator Cluster Survey; DHS=Demographic Health Survey; FGM/C: female genital mutilation; SD: standard deviation; HIECS: Household Income, Expenditure and Consumption Surveys; DV: Domestic violence; CTS-PC=Parent–Child Conflict Tactics Scale; AIDS=acquired immunodeficiency syndrome; ICAST= Standard Child Abuse Screening Tool; CT = Children's Institutional Version; NTI= National Tuberculosis Institute; PTB=pulmonary tuberculosis; BDI= Beck Depression Inventory; FV=family violence; HBMNC = Home Based Maternal and Neonatal Care; SDQ = Strengths and Difficulties Questionnaire; WLWH = women living with HIV; SRHR = sexual and reproductive health and rights; PLWH = people living with HIV
