## Supplementary material for "Violence against women and children in Yemen: A mixed-methods systematic review": S5. Table. Quality assessment

### S5 Table. Quality assessment- Mixed Method Analysis Tool (MMAT) [1]

| **Author, year with publication** | **Screening** | | **Qualitative** | | | | | **Quantitative** | | | | | | | | | | | | | | | **Mixed methods** | | | | |
| --- | --- | --- | --- | --- | --- | --- | --- | --- | --- | --- | --- | --- | --- | --- | --- | --- | --- | --- | --- | --- | --- | --- | --- | --- | --- | --- | --- |
|  |  |  |  |  |  |  |  | **Quant. Randomized** | | | | | **Quant Non-Randomized** | | | | | **Descriptive** | | | | |  |  |  |  |  |
|  | **A** | **B** | **Q1** | **Q2** | **Q3** | **Q4** | **Q5** | **Q6** | **Q7** | **Q8** | **Q9** | **Q10** | **Q11** | **Q12** | **Q13** | **Q14** | **Q15** | **Q16** | **Q17** | **Q18** | **Q19** | **Q20** | **Q21** | **Q22** | **Q23** | **Q24** | **Q25** |
| Akmatov, 2011  [2] | Y | Y | - | - | - | - | - | - | - | - | - | - | - | - | - | - | - | Y | Y | Y | Y | Y | - | - | - | - | - |
| Al Taj, 2023 [3] | Y | Y | - | - | - | - | - | - | - | - | - | - | - | - | - | - | - | Y | Y | Y | Y | Y | - | - | - | - | - |
| Alansari, 2006 [4] | Y | Y | - | - | - | - | - | - | - | - | - | - | - | - | - | - | - | C | Y | Y | C | Y | - | - | - | - | - |
| Alizzy, 2017 [5] | Y | Y | - | - | - | - | - | - | - | - | - | - | - | - | - | - | - | Y | Y | Y | C | Y | - | - | - | - | - |
| Al-Khulaidi, 2013 [6] | Y | Y | - | - | - | - | - | - | - | - | - | - | - | - | - | - | - | Y | Y | Y | Y | Y | - | - | - | - | - |
| Alosaimi, 2019 [7] | Y | Y | - | - | - | - | - | - | - | - | - | - | - | - | - | - | - | Y | Y | Y | Y | Y | - | - | - | - | - |
| Alyahri, 2008 [8] | Y | Y | - | - | - | - | - | - | - | - | - | - | - | - | - | - | - | C | Y | Y | Y | Y | - | - | - | - | - |
| Assaad, 2019 [9] | Y | Y | - | - | - | - | - | - | - | - | - | - | - | - | - | - | - | Y | Y | Y | C | Y | - | - | - | - | - |
| Badahdah, 2016 [10] | Y | Y | - | - | - | - | - | - | - | - | - | - | - | - | - | - | - | C | N | C | Y | Y | - | - | - | - | - |
| Bamatraf, 2019 [11] | Y | Y | - | - | - | - | - | - | - | - | - | - | - | - | - | - | - | Y | Y | Y | Y | Y | - | - | - | - | - |
| Ba-Obaid, 2002 [12] | Y | Y | Y | Y | Y | Y | Y | - | - | - | - | - | - | - | - | - | - | - | - | - | - | - | Y | Y | Y | C | Y |
| Ba-Saddik, 2012 [13] | Y | Y | - | - | - | - | - | - | - | - | - | - | - | - | - | - | - | Y | Y | Y | Y | Y | - | - | - | - | - |
| Ba-Saddik, 2013 [14] | Y | Y | - | - | - | - | - | - | - | - | - | - | - | - | - | - | - | Y | Y | Y | Y | Y | - | - | - | - | - |
| Chamberlain, 2007 [15] | Y | Y | - | - | - | - | - | - | - | - | - | - | - | - | - | - | - | Y | Y | Y | C | Y | - | - | - | - | - |
| Christiansen, 2019 [16] | N | N | N | N | Y | C | C | - | - | - | - | - | - | - | - | - | - | - | - | - | - | - | - | - | - | - | - |
| Costa, 2017 [17] | Y | Y | - | - | - | - | - | - | - | - | - | - | - | - | - | - | - | C | Y | Y | Y | Y | - | - | - | - | - |
| Date, 2005 [18] | Y | Y | - | - | - | - | - | - | - | - | - | - | - | - | - | - | - | Y | C | Y | Y | C | - | - | - | - | - |
| El Tantawi, 2018 [19] | Y | Y | - | - | - | - | - | - | - | - | - | - | - | - | - | - | - | C | C | C | C | Y | - | - | - | - | - |
| ICRW & UNICEF, 2017 [20] | Y | Y | Y | Y | Y | Y | Y | - | - | - | - | - | - | - | - | - | - | - | - | - | - | - | - | - | - | - | - |
| Kempe, 2015 [21] | Y | Y | Y | Y | Y | Y | Y | - | - | - | - | - | - | - | - | - | - | - | - | - | - | - | - | - | - | - | - |
| Lewis, 2012 [22] | Y | Y | Y | Y | Y | C | Y | - | - | - | - | - | - | - | - | - | - | - | - | - | - | - | Y | Y | Y | C | Y |
| Ouis, 2009 [23] | Y | Y | Y | Y | Y | Y | Y | - | - | - | - | - | - | - | - | - | - | - | - | - | - | - | - | - | - | - | - |
| Sharaf , 2019 [24] | Y | Y | - | - | - | - | - | - | - | - | - | - | - | - | - | - | - | Y | Y | Y | C | Y | - | - | - | - | - |
| Smits, 2013 [25] | Y | Y | - | - | - | - | - | - | - | - | - | - | - | - | - | - | - | Y | Y | Y | Y | Y | - | - | - | - | - |
| Yoder, 2013 [26] | Y | Y | - | - | - | - | - | - | - | - | - | - | - | - | - | - | - | Y | Y | Y | Y | Y | - | - | - | - | - |

**Legend**: Y: yes; N: no; C: can’t tell.; (-) NA (Quantitative randomized controlled trials & Quantitative nonrandomized: Questions not presented as these studies are “na”

Screening questions (for all types)

A: Are there clear research questions?

B: Do the collected data allow to address the research questions?

Qualitative

Q1: Is the qualitative approach appropriate to answer the research question?

Q2: Are the qualitative data collection methods adequate to address the research question?

Q3: Are the findings adequately derived from the data?

Q4: Is the interpretation of results sufficiently substantiated by data?

Q5: Is there coherence between qualitative data sources, collection, analysis, and interpretation?

Quantitative descriptive

Q16: Is the sampling strategy relevant to address the research question?

Q17: Is the sample representative of the target population?

Q18: Are the measurements appropriate?

Q19: Is the risk of nonresponse bias low?

Q20: Is the statistical analysis appropriate to answer the research question?

Mixed methods

Q21: Is there an adequate rationale for using a mixed methods design to address the research question?

Q22: Are the different components of the study effectively integrated to answer the research question?

Q23: Are the outputs of the integration of qualitative and quantitative components adequately interpreted?

Q24: Are divergences and inconsistencies between quantitative and qualitative results adequately addressed?

Q25: Do the different components of the study adhere to the quality criteria of each tradition of the methods involved
